# Quantitative genome-scale analysis of human liver reveals dysregulation of glycosphingolipid pathways in progressive nonalcoholic fatty liver disease

**DOI:** 10.1101/2021.02.09.21251354

**Authors:** Partho Sen, Olivier Govaere, Tim Sinioja, Aidan McGlinchey, Dawei Geng, Vlad Ratziu, Elisabetta Bugianesi, Jörn M. Schattenberg, Antonio Vidal-Puig, Michael Allison, Simon Cockell, Ann K. Daly, Tuulia Hyötyläinen, Quentin M. Anstee, Matej Orešič

**Affiliations:** School of Medical Sciences, Örebro University, 70281 Örebro, Sweden; Turku Bioscience Centre, University of Turku and Åbo Akademi University, FI-20520 Turku, Finland; Translational & Clinical Research Institute, Faculty of Medical Sciences, Newcastle University, Newcastle upon Tyne, NE2 4HH, UK; Department of Chemistry, Örebro University, 70281 Örebro, Sweden; Assistance Publique-Hôpitaux de Paris, hôpital Beaujon, University Paris-Diderot, Paris, France; Department of Medical Sciences, Division of Gastro-Hepatology, A.O. Città della Salute e della Scienza di Torino, University of Turin, Turin, Italy; Metabolic Liver Research Programm, Department of Medicine, University Hospital Mainz, Mainz, Germany; University of Cambridge Metabolic Research Laboratories, Wellcome-MRC Institute of Metabolic Science, Addenbrooke’s Hospital, Cambridge CB2 0QQ, UK; Liver Unit, Department of Medicine, Cambridge Biomedical Research Centre, Cambridge University NHS Foundation Trust, United Kingdom; Newcastle NIHR Biomedical Research Center, Newcastle upon Tyne Hospitals NHS Foundation Trust, Newcastle upon Tyne, UK

## Abstract

Nonalcoholic fatty liver disease (NAFLD) is an increasingly prevalent disease that is associated with multiple metabolic disturbances, yet the metabolic pathways underlying its progression are poorly understood. Here we studied metabolic pathways of the human liver across the full histological spectrum of NAFLD. We analyzed whole liver tissue transcriptomics and serum metabolomics data obtained from a large, prospectively enrolled cohort of 206 histologically characterized patients derived from the European NAFLD Registry and developed genome-scale metabolic models (GEMs) of human hepatocytes at different stages of NAFLD. We identified several metabolic signatures in the liver and blood of these patients, specifically highlighting the alteration of vitamins (A, E) and glycosphingolipids, and their link with complex glycosaminoglycans in advanced fibrosis. Furthermore, we derived GEMs and identified metabolic signatures of three common NAFLD-associated gene variants *(PNPLA3, TM6SF2* and *HSD17B13)*. The study demonstrates dysregulated liver metabolic pathways which may contribute to the progression of NAFLD.

## Introduction

Nonalcoholic fatty liver disease (NAFLD) represents a histological spectrum characterized by increased lipid accumulation in the hepatocytes, that encompasses non-alcoholic fatty liver (‘simple’ steatosis, NAFL) and an inflammatory form termed as nonalcoholic steatohepatitis (NASH) in which progressive hepatic fibrosis occurs, ultimately leading to cirrhosis and hepatocellular carcinoma (HCC) in some patients (Anstee et al., 2019). NAFLD is associated with features of the metabolic syndrome including obesity, type 2 diabetes mellitus (T2DM), dyslipidemia, and hypertension (Anstee et al., 2013; Labenz et al., 2018). The global prevalence of NAFLD in the general population has been estimated to be 25%, whilst the prevalence of NASH has been estimated to range from 3 – 5% (Younossi et al., 2018). The prevalence of NAFLD has been increasing proportionately with the epidemics of obesity and is predicted to continue to rise (Estes et al., 2018).

The natural history of NAFLD is highly variable, with substantial interpatient variation in disease severity and outcome predicted by the degree of fibrosis (McPherson et al., 2015; Taylor et al., 2020). NAFLD is generally considered to be a complex disease trait driven by an obesogenic environment acting on a background of genetic susceptibility (Anstee et al., 2020). However, knowledge about the processes that contribute to the observed variation in severity remains incomplete. There is a pressing need to elucidate the pathophysiological processes that occur as NAFL progresses to NASH and fibrosis stage increases towards cirrhosis. Such knowledge will define novel biomarkers that better risk stratify patients, helping to individualize their care, and aid the development of novel therapeutic strategies and drug targets.

Progression of NAFLD is characterized by distinct metabolic changes in the liver (Masoodi et al., 2021, in press). Previously, we showed that there is an excess accumulation of liver triacylglycerols (TGs) in NAFLD, particularly those with low carbon number and double bond content (Oresic et al., 2013), which reflects increased *de novo* lipogenesis in NAFLD (Kotronen et al., 2009; Westerbacka et al., 2010).

Over the past decade, genome-scale metabolic modeling (GSMM) has emerged as a powerful tool to study metabolism in human cells (Bordbar et al., 2012; Sen et al., 2020; Sen et al., 2017), including in modeling metabolism of human liver under healthy and disease conditions (Hyotylainen et al., 2016; Jerby et al., 2010; Mardinoglu et al., 2014). In a previous study, we charted metabolic activities associated with degree of steatosis in human liver from 16 NAFLD cases by integrating genome-wide transcriptomics data from human liver biopsies, and metabolic flux data measured across the human splanchnic vascular bed using GSMM (Hyotylainen et al., 2016). A similar GSMM study in 32 patients with NAFLD and 20 controls identified serine deficiency and suggested that the serum concentrations of glycosaminoglycans (GAGs) (*e*.*g*., chondroitin and heparan sulphates) associate with the staging of NAFLD (Mardinoglu et al., 2014). Both studies, whilst informative, only used transcriptomic data from relatively small patient cohorts, with crude histological characterization of disease severity, thus neither study adequately addressed grade of steatohepatitis or stage of fibrosis with sufficient granularity. In addition, several other studies in human and mice have identified molecular signatures in the liver associated with NAFLD (Arendt et al., 2015; Haas et al., 2019; Lefebvre et al., 2017; Moylan et al., 2014; Starmann et al., 2012). Moylan *et al*., compared the hepatic gene expression profiles in high-(severe) and low-risk (mild) NAFLD patients and idenfitied specific metabolic pathways that were differentially activated in these groups (Moylan et al., 2014). In a cross-sectional study, Arendt *et al*., compared the hepatic gene expression in subjects with either healthy liver, NAFL or NASH and showed marked alteration in the gene expression profiles associated with the polyunsaturated fatty acid metabolism (Arendt et al., 2015). Integrative analyisis of human transcriptomic datasets and the mice models have identified functional role of dermatopontin in collagen deposition and hepatic fibrosis (Lefebvre et al., 2017). Futhermore, transcriptomic analysis and immune profiling of NASH patients also relvealed that several genes associated with the inflammatory responses were altered in progression to NASH (Haas et al., 2019). However, these studies have largely employed microarray-based techniques and hence, lacked the comprehensive approach provided by the genome-wide RNA sequencing (RNA-seq) (Govaere et al., 2020).

Herein, we examined the changes in hepatic metabolic processes and pathways that occur across the full histological spectrum of NAFLD, exploring the evolving changes during the progression of NAFL to NASH with increasing fibrosis stages including cirrhosis (F0 up to F4). Whole liver tissue RNA-Seq transcriptomic data from a cohort of histologically characterized patients derived from the European NAFLD Registry (n=206) (Govaere et al., 2020; Hardy et al., 2020) was used to develop personalized and NAFLD-stage-specific genome-scale metabolic models (GEMs) of human hepatocytes (**Table 1** and **Fig 1**). The integrative approach employed in this study defined the changes of liver metabolism in NAFL and with progressive NASH-associated fibrosis. Furthermore, GSMM predicted the metabolic differences among carriers of widely validated genetic variants associated with NAFLD / NASH disease severity in three genes *(PNPLA3, TM6SF2* and *HSD17B13)* (Abul-Husn et al., 2018; Anstee et al., 2020; Liu et al., 2014; Romeo et al., 2008).

**Table 1.**
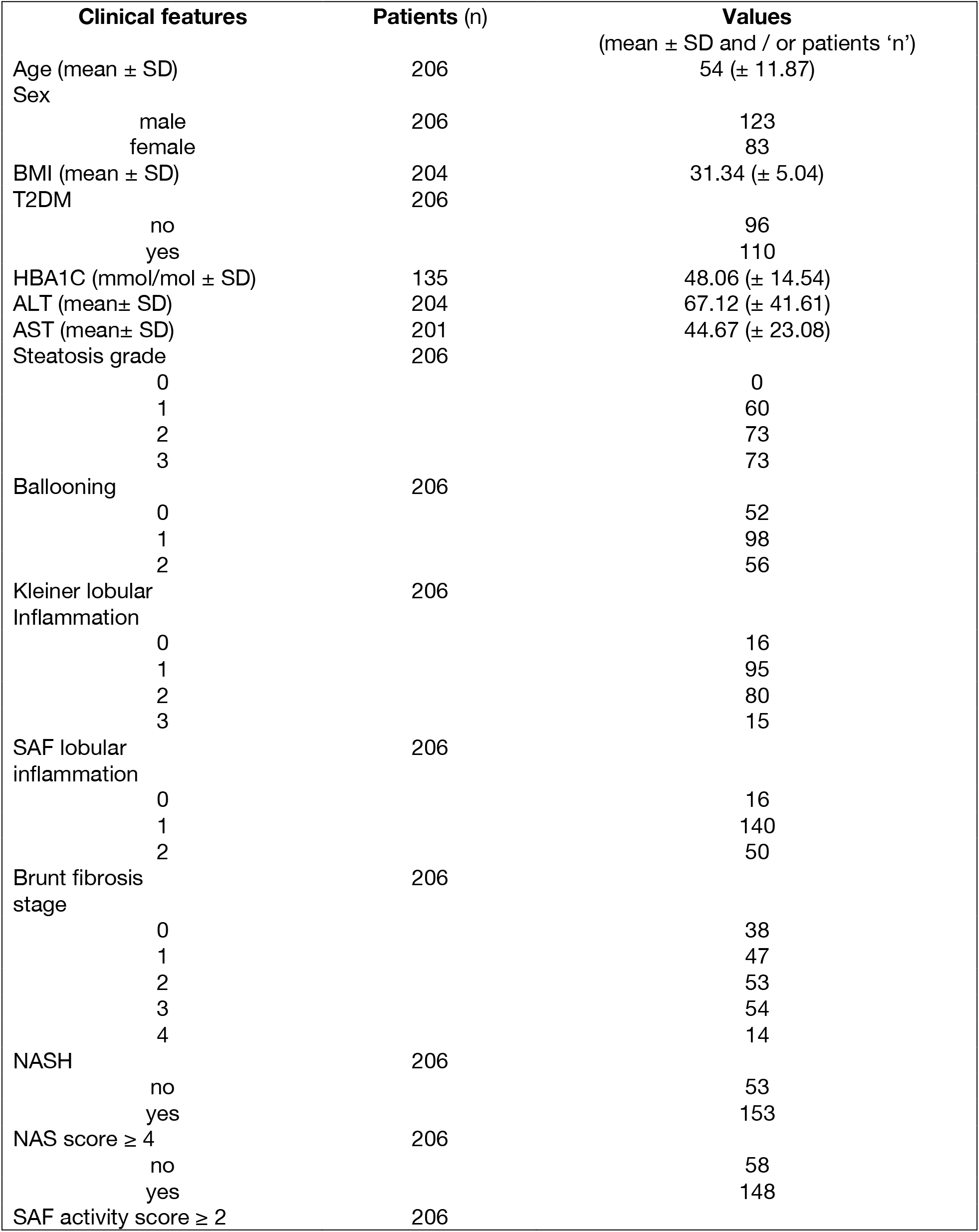
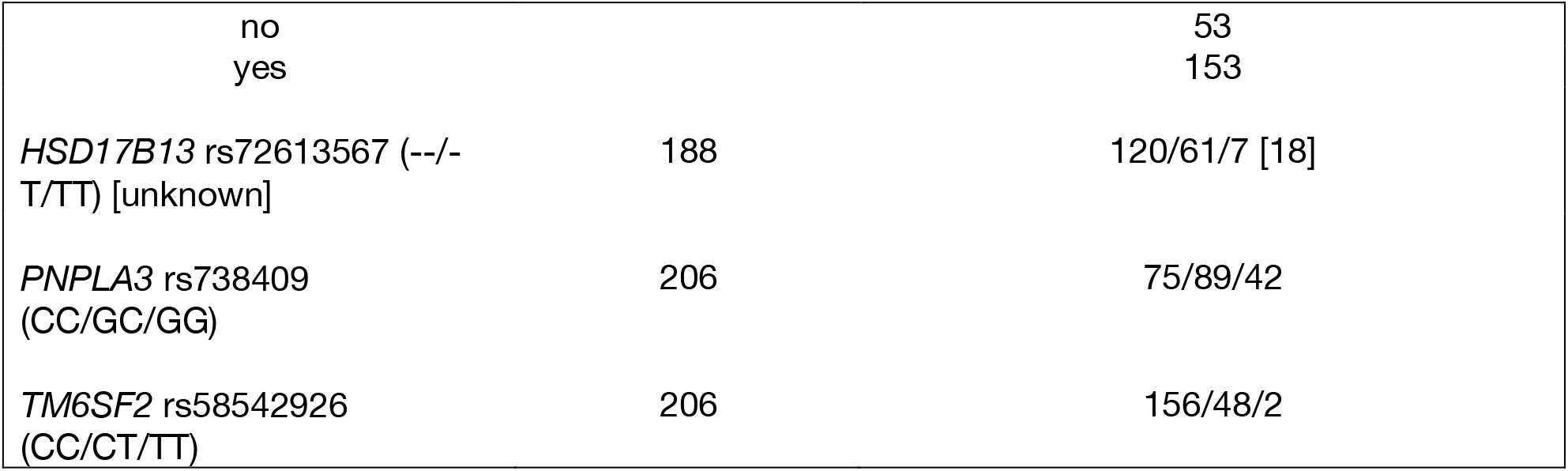
Demographic characteristics of the EPoS RNA-Seq Cohort.

| **Clinical features** | **Patients (n)** | **Values**<br>(mean ± SD and / or patients 'n') |
| --- | --- | --- |
| Age (mean ± SD) | 206 | 54 (± 11.87) |
| Sex |  |  |
| male | 206 | 123 |
| female |  | 83 |
| BMI (mean ± SD) | 204 | 31.34 (± 5.04) |
| T2DM | 206 |  |
| no |  | 96 |
| yes |  | 110 |
| HBA1C (mmol/mol ± SD) | 135 | 48.06 (± 14.54) |
| ALT (mean ± SD) | 204 | 67.12 (± 41.61) |
| AST (mean ± SD) | 201 | 44.67 (± 23.08) |
| Steatosis grade | 206 |  |
| 0 |  | 0 |
| 1 |  | 60 |
| 2 |  | 73 |
| 3 |  | 73 |
| Ballooning | 206 |  |
| 0 |  | 52 |
| 1 |  | 98 |
| 2 |  | 56 |
| Kleiner lobular<br>Inflammation | 206 |  |
| 0 |  | 16 |
| 1 |  | 95 |
| 2 |  | 80 |
| 3 |  | 15 |
| SAF lobular<br>inflammation | 206 |  |
| 0 |  | 16 |
| 1 |  | 140 |
| 2 |  | 50 |
| Brunt fibrosis<br>stage | 206 |  |
| 0 |  | 38 |
| 1 |  | 47 |
| 2 |  | 53 |
| 3 |  | 54 |
| 4 |  | 14 |
| NASH | 206 |  |
| no |  | 53 |
| yes |  | 153 |
| NAS score ≥ 4 | 206 | |
| no |  | 58 |
| yes |  | 148 |
| SAF activity score ≥ 2 | 206 | |

| no |  | 53 |
| --- | --- | --- |
| yes |  | 153 |
| *HSD17B13* rs72613567 (--/-<br>T/TT) [unknown] | 188 | 120/61/7 [18] |
| *PNPLA3* rs738409<br>(CC/GC/GG) | 206 | 75/89/42 |
| *TM6SF2* rs58542926<br>(CC/CT/TT) | 206 | 156/48/2 |

**Figure 1.**
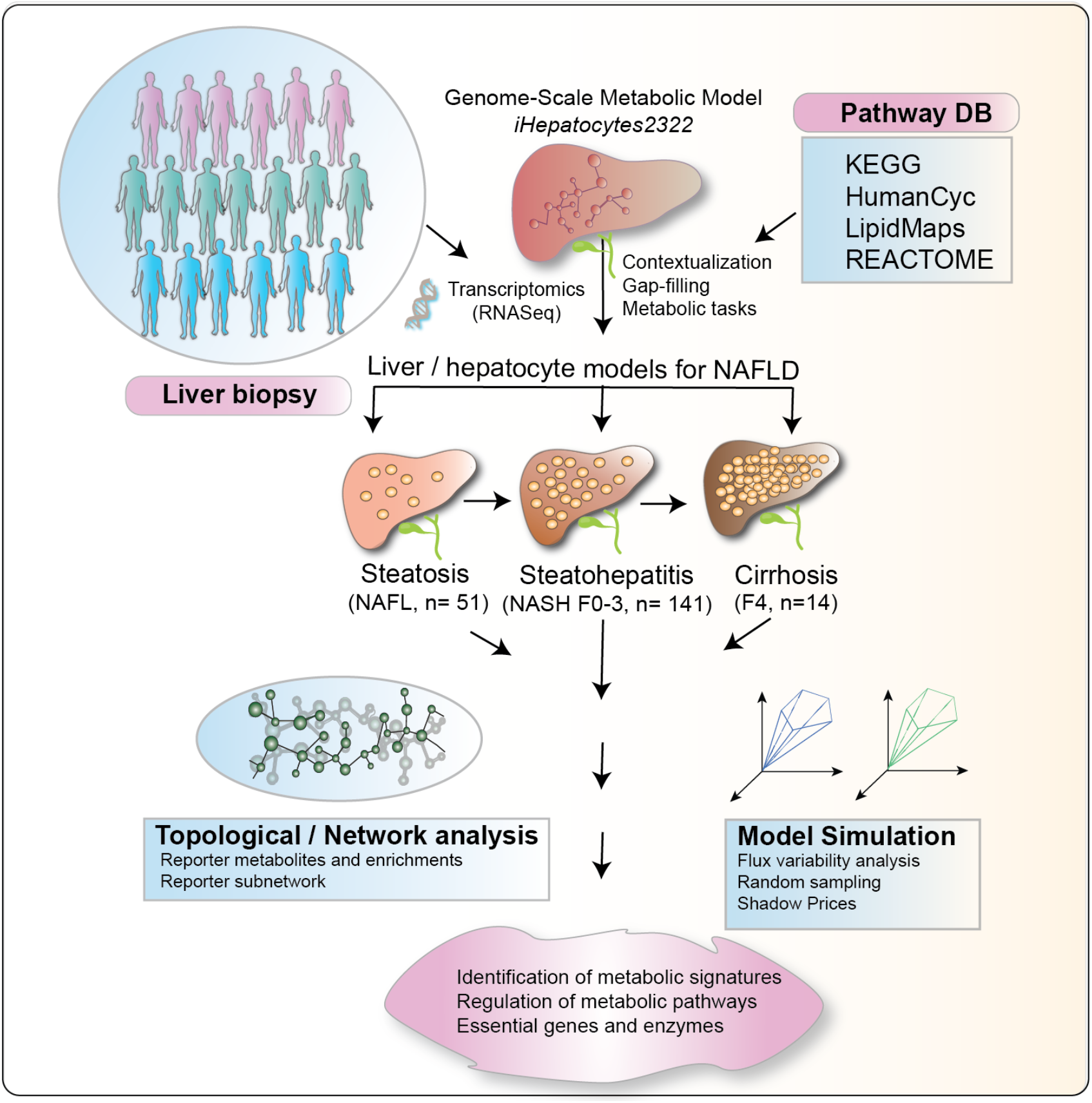
Study design and schematic illustration. Key steps in the reconstruction of stage-specific genome-scale metabolic models (GEMs) of human hepatocytes in NAFLD. Labels F(0-4) represents different stages of fibrosis.

## Results

### Development of genome-scale metabolic models of human hepatocytes in the patients with NAFLD

A GEM of human hepatocyte (*iHepatocytes2322*) was first developed by Mardinoglu *et al*., by combining the *iHepatocyte1154* GEM (Agren et al., 2012) with previously published liver models. *iHepatocytes2322* comprises 2,322 genes, 5,686 metabolites and 7,930 reactions found in the human liver (Mardinoglu et al., 2014). Here, by using NAFLD stage-specific transcriptomics data generated from the liver biopsies of 206 patients with NAFLD (**Table 1**) (Govaere et al., 2020), we contextualized the *iHepatocytes2322* model and developed personalized GEMs of human hepatocytes across the full spectrum of NAFLD severity (NAFL through NASH F0-F4) (**Fig 1**). A stepwise strategy that combines *iMAT* (Zur et al., 2010) and *E-*flux (Colijn et al., 2009) algorithm was developed for the contextualization of GEMs, *i*.*e*., selection of stage-specific active reactions and their associated genes and metabolites from the *iHepatocytes2322*. The number of genes, metabolites and reactions included in these models are given in **Fig S1**. All the models were tested to carry out 256 metabolic tasks as given in (Mardinoglu et al., 2013; Mardinoglu et al., 2014). The occurrence scores of the metabolic tasks carried out by these models are shown in **Fig S2**.

The hepatocyte GEMs were used to study the metabolic differences across the NAFLD spectrum. To increase the statistical power of differential expression analysis of the transcriptomic data required for GSMM, cases were grouped by disease severity and clinical implications for outcome (Taylor et al., 2020): ‘minimal’ disease – NAFL + NASH (F0-1) (n= 85); ‘mild’ disease – NAFL + NASH (F0-2) (n= 138); ‘clinically significant’ non-cirrhotic fibrosis – (NASH F2-3) (n= 107); and ‘advanced’ fibrosis – (NASH F3-4) (n= 68).

### Stage-specific alterations of the metabolic pathways at various stages of NAFLD

Partial least squares-discriminant analysis (PLS-DA) (Le Cao et al., 2011) of the metabolic fluxes predicted by the personalized hepatocyte GEMs showed that patients with NASH (F3, AUC=0.70) and cirrhosis (F4, AUC=0.83) were metabolically different from NAFL and NASH (F0-1 and F2) groups (**Fig 2A** and **2C**). Here, the reaction flux states were estimated by a non-uniform random sampling method that finds solutions among the feasible flux distributions of the metabolic network (Bordel et al., 2010; Osterlund et al., 2013). The PLS-DA model has identified several metabolic subsystems/pathways such as fatty acid, glycerophspholipid, cholesterol, bile acid, tryptophan, histidine, glutathione, vitamin (B,D), nucleotide metabolism and ROS detoxification which were altered (Variable Importance in Projection (VIP) scores (Farrés et al., 2015) >1) among these groups (**Fig 2B**). Pairwise comparisons between different NAFLD groups suggested that most of these subsystems were upregulated in the patients with ‘clinically significant’ non-cirrhotic or ‘advanced’ fibrosis as compared to ‘mild’ or ‘minimal’ diseases (**Fig S3**). Intriguingly, at ‘advanced’ (*vs*. ‘mild’) fibrosis, fluxes across the sphingolipid metabolism were significantly (two sample t-test, p-values < 0.05, corrected for False Discovery Rate (FDR)) upregulated, whilst glycosphingolipid (GSL) biosynthesis–globo series were downregulated (**Fig S3**).

**Figure 2.**
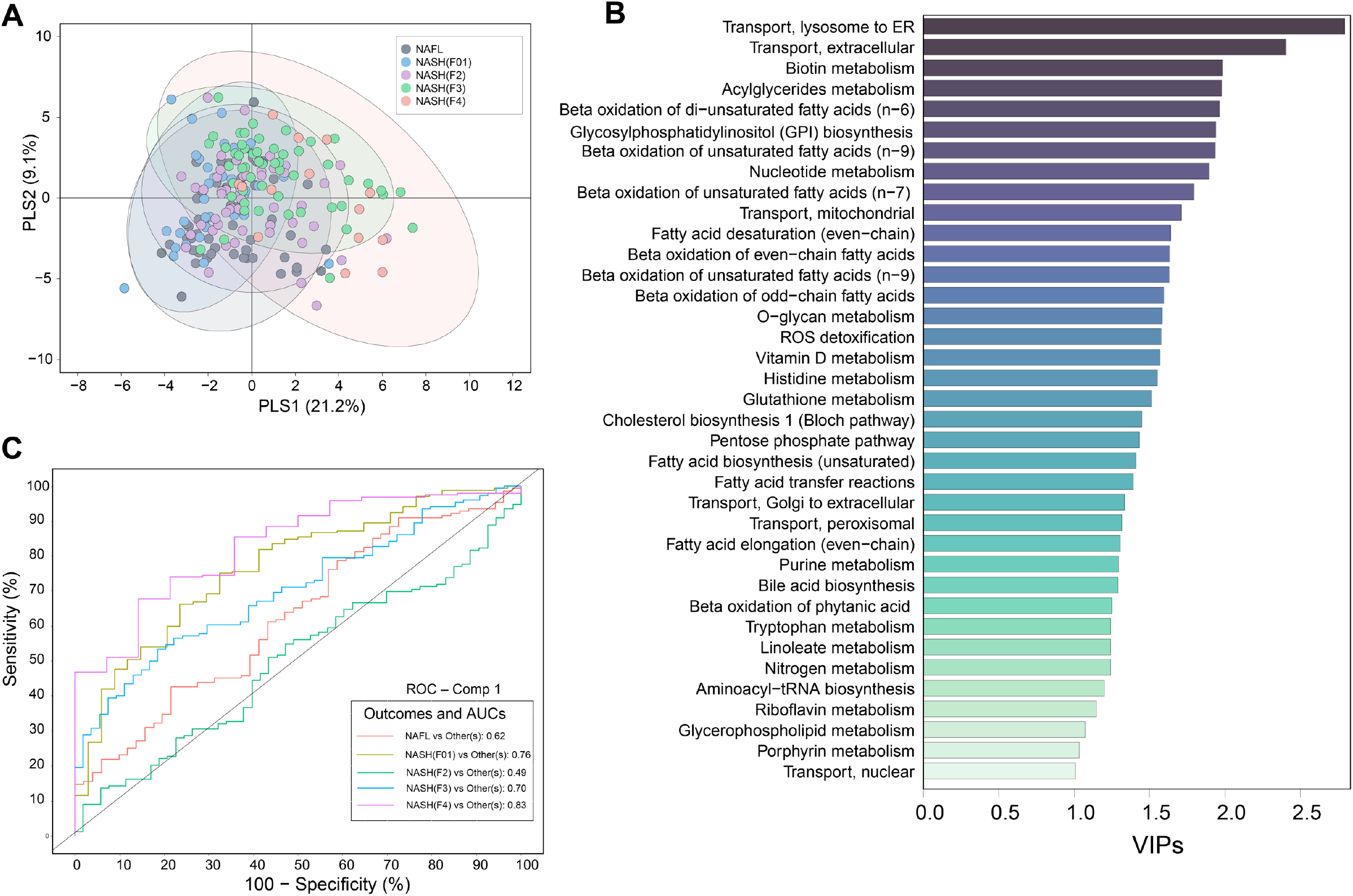
Metabolic pathways along the NAFLD spectrum. (**A**) PLS-DA–score plot showing metabolic pathway differences across NAFL and NASH (F0-4) patient groups. (**B**) Metabolic pathways/subsystems with (PLS-DA, VIP score >1). (**C**) Class-specific AUCs and ROC curves of the PLS-DA model.

### Reporter metabolite analysis of human hepatocytes unveiled the intrinsic regulation of GSLs, GAGs and cholesterols in advance fibrosis

Reporter metabolite (RM) analysis is a metabolite-centric differential approach that aids in identifying metabolites in a network around which significant transcriptional changes occur (Cakir et al., 2006; Patil and Nielsen, 2005). RMs can predict hot spots in a metabolic network that are significantly altered between two different conditions, in this case, different stages of NAFLD.

RM analysis revealed that several species of eicosanoids, estrone, steroids and retinoic acids were upregulated (p < 0.05, corrected for FDR) in NASH F2 stage as compared to NAFL (**Fig S4**). RMs of NASH F3 (*vs*. NAFL) showed a remarkable pattern with several species of GSLs, particularly, cerebrosides (glucosylceramides (GlcCers), lactosylceramides (LacCers), digalactosylceramides), globosides and gangliosides (GM1-, GM2-alpha) were downregulated (p < 0.05, corrected for FDR). At this stage, RMs of GAGs (heparin, keratin sulphates) and cholesterols were also downregulated (p < 0.05, corrected for FDR) (**Fig 3**). Intriguingly, these GAGs were linked to GSLs by 3’-Phosphoadenosine-5’-phosphosulfate (PAPs) and globotriaosylceramides which in turn were also downregulated (p < 0.05, corrected for FDR) (**Fig 3**). In addition, RMs of multiple eicosanoids, prostaglandins (PGs), leukotrienes (LTs) and arachidonic and retinoic acid derivates were upregulated downregulated (p < 0.05, corrected for FDR) (**Fig 3**).

**Figure 3.**
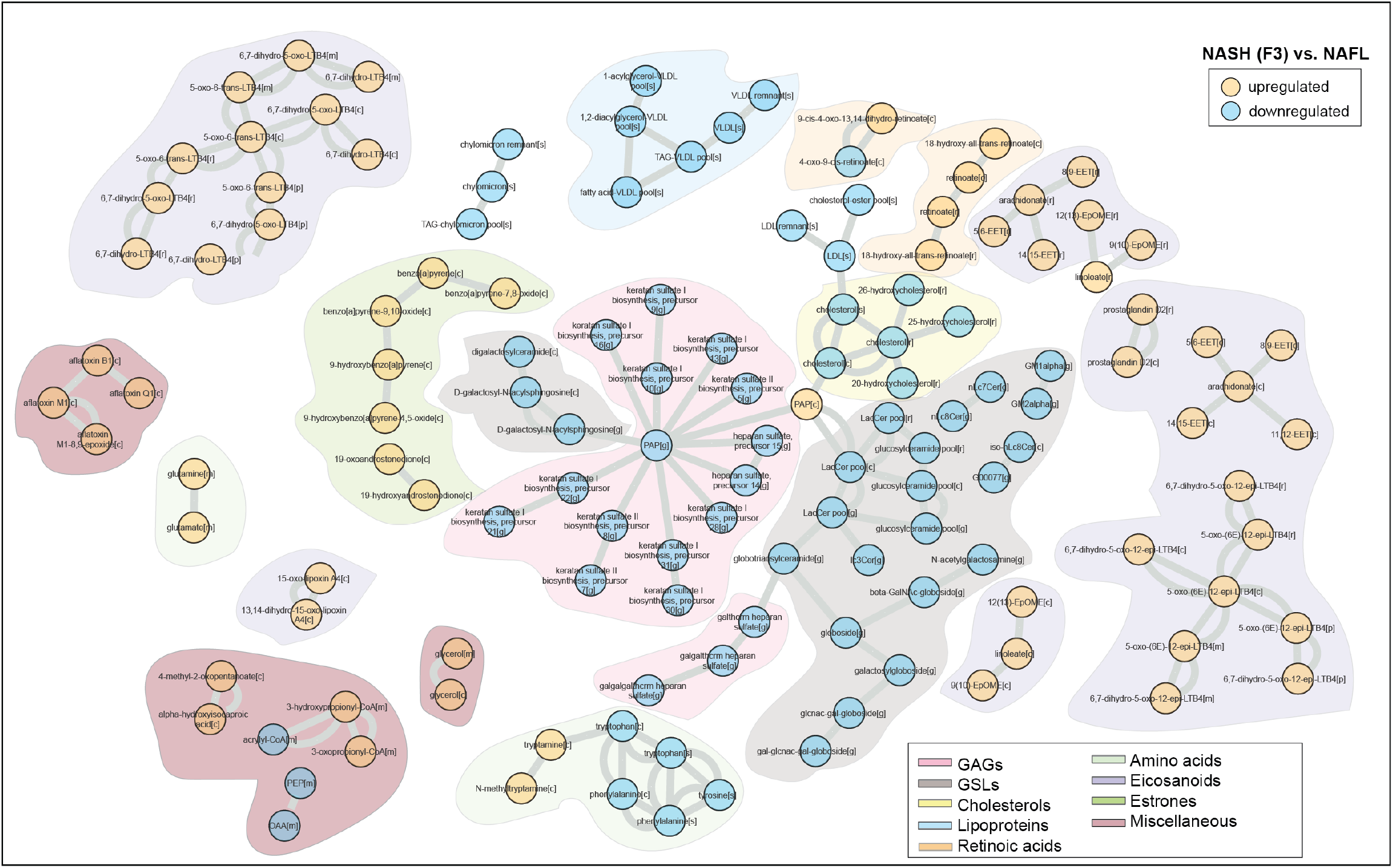
Reporter metabolites of NASH F3 *vs*. NAFL. A metabolic-centric view of reporter metabolite (RM) modules that were significantly altered (p<0.05) between NASH F3 *vs*. NAFL. Orange and blue color denotes up and downregulated respectively. Each node represents a ‘RM’ and single or double lines represent reversible or irreversible metabolic reactions respectively. RMs that belong to a particular chemical class are color coded.

Interestingly, RMs of GSLs, GAGs, and cholesterols remain downregulated (p < 0.05, corrected for FDR) in cirrhosis (NASH F4 *vs*. NAFL) (**Fig S5**). A similar directional change in the RMs of GSLs, GAGs, cholesterols, gamma-tocopherols (Vitamin E) was observed in ‘advanced’ fibrosis – (NASH F3-4) vs. ‘minimal disease – NAFL + NASH (F0-1) (**Fig S6**) and ‘mild disease’ – NAFL + NASH (F0-2), respectively (**Fig S7**). In addition, RMs of aromatic and sulphur containing amino acids, *i*.*e*., phenylalanine, tyrosine, tryptophan, histidine and methionine were increased in the ‘advanced’ fibrosis vs. ‘mild disease’. These amino acids were associated with apolipoproteins (apo), primarily involved in the formation of low-density lipoprotein particles (LDL and VLDL). However, no significant change in the GSLs, GAGs, cholesterols was observed in the ‘clinically significant’ non-cirrhotic fibrosis – (NASH F2-3) vs. ‘minimal disease’ (**Fig S8** and **S9**).

### Regulation of sphingolipid and ceramide pathways in advanced fibrosis

GSLs and ceramides (Cers) are essential intermediates of sphingolipid (SL) pathways (Sen et al., 2021) (**Fig 4A**). They play a significant role in maintaining the integrity of the plasma membrane. Our results suggest that, several species of GSLs and related pathways were altered at NASH (F3 and F4) stages (**Fig 3, S5** and **S9**). At these stages, the GSLs were associated with GAGs (heparin, keratin sulphates) and cholesterols which was also altered (**Fig 3** and **S9**). Therefore, the intracellular regulation of SL pathways might play a critical role in the progression of fibrosis.

**Figure 4.**
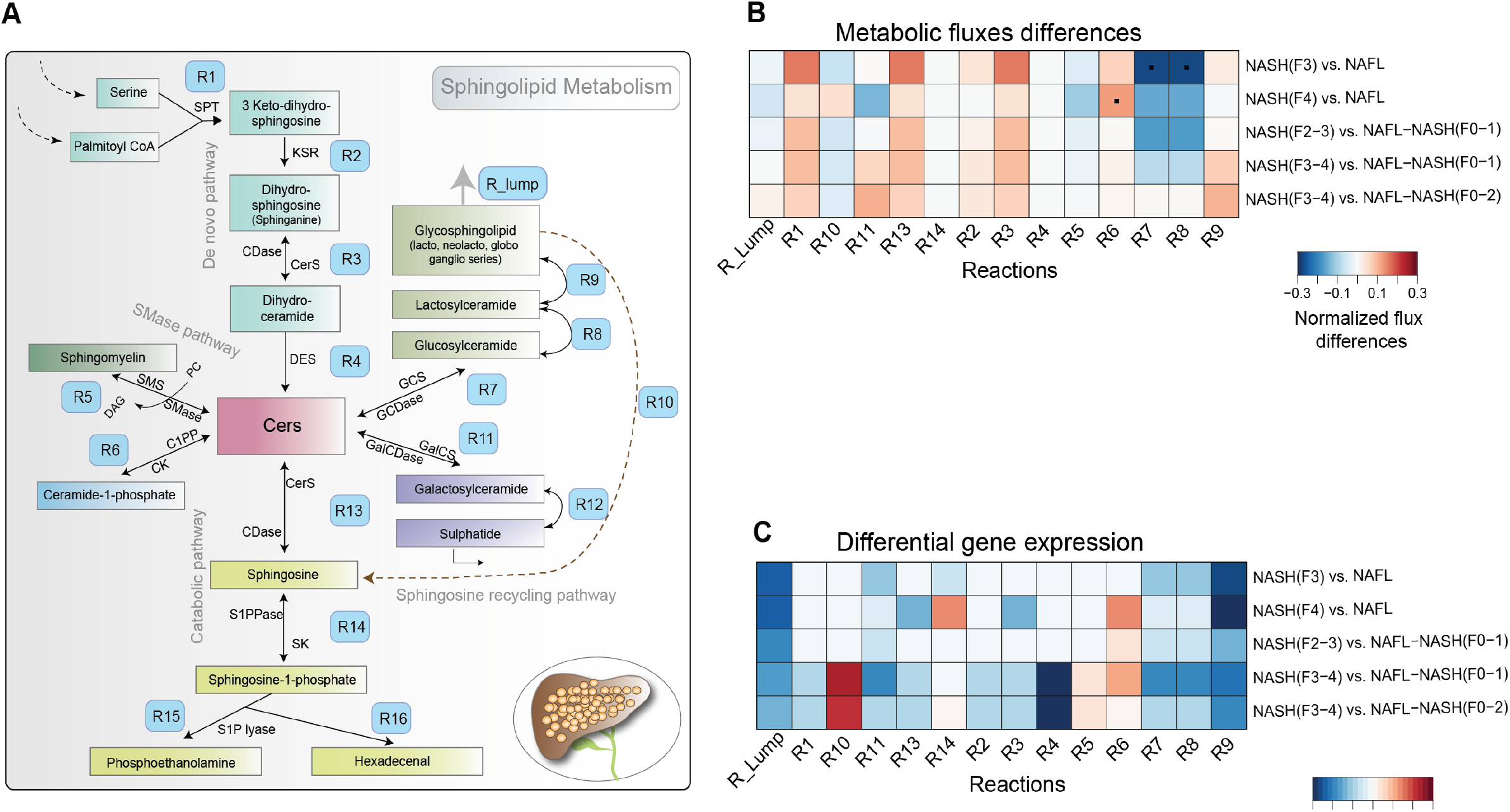
Regulation of sphingolipid metabolism in NAFLD. (**A**) A schematic illustration of sphingolipid (SL) metabolism in human hepatocytes. Abbreviations: *SPT, serine palmitoyl-CoA transferase; CerS, ceramide synthase; and DES, dihydroceramide desaturase; SMase, sphingomyelinase pathway S1P, sphingosine-1-phosphate; CDase, ceramidases; SK, sphingosine kinase; S1PPase, sphingosine phosphate phosphatases GCDase, glucosidase; GalCDase, galactosidase; C1P, ceramide 1-phosphate*. The modeled reactions are labeled with ‘R#’. (**B**) Averaged normalized flux differences across the SL reactions of the hepatocytes at different stages of NAFLD. Red and blue color denotes up- and downregulation of reaction fluxes within at two different stages of NAFLD. Reaction fluxes at a particular NAFLD stage was estimated by using a non-uniform random sampling method that finds solutions among the feasible flux distributions of the metabolic network. *p<0.05 (two-sample t-test, corrected for FDR) (**C**) Log_2_ fold change (FC) of the differentially expressed (p<0.05 corrected for FDR) metabolic genes (MGs) of SL pathways at two different stages of NAFLD. Expression of multiple genes mapped to a particular reaction was determined by GPR rules.

**Figure 5.**
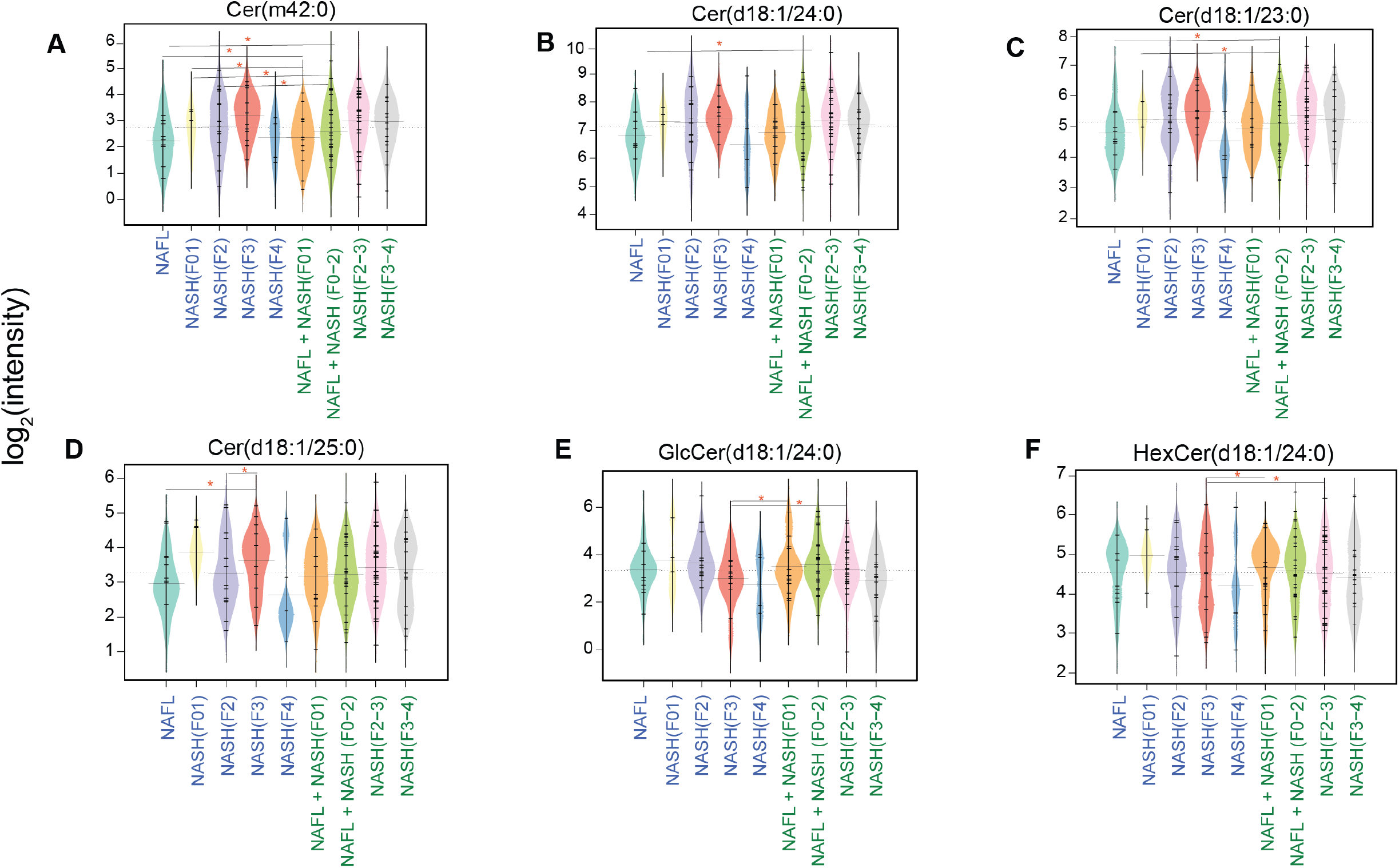
Serum metabolic levels of ceramides, glycosphingolipids in the patients at different stages of NAFLD. Beanplots showing the log_2_ intensities of the metabolites across different stages of NAFLD. The dotted line denotes the mean of the population, and the black dashed lines in the bean plots represent the group mean. (**A–D**) Ceramides, (**E-F**) Glucosyl-(GlcCers) and Hexosylceramides (HexCers). *p<0.05 (Tukey’s HSD)

Next, we identified the metabolic genes (MGs) of the sphingolipid (SL) pathways that were differentially expressed (p < 0.05, corrected for FDR) with the progression of NASH-associated fibrosis. Intriguingly, MGs and fluxes of glycosyl-(GlcCer) and lactosylceramide (LacCer) biosynthesis pathways (R7, R8, R9) were downregulated in ‘advanced’ fibrosis *vs*. ‘minimal’ and/or ‘mild’ disease stages (**Fig 4A-C**). Moreover, the net flux and gene expression of (R6) reaction was upregulated (p < 0.05, corrected for FDR) in ‘advanced’ fibrosis *vs*. ‘mild disease which suggests conversion of Cer to bioactive ceramide-1-phosphate (**Fig 4A-C**).

Taken together, in ‘advanced’ fibrosis the hepatocytes tend to decreases the synthesis of GSLs, particularly GlcCers, LacCers and GalCers from Cers (**Fig 4A-C**). Excessive accumulation of Cers in the hepatocyte may be required to exert cellular signaling, modulate inflammatory response or generate apoptotic stress signals (Apostolidis et al., 2016; Zhang et al., 2019).

### Marked alterations of Cers and GSLs levels in the serum of the patients with advanced fibrosis

By applying lipidomics to serum samples obtained from NAFLD patients in the present study, we measured the levels of Cers and GSLs. We found that the serum levels of several Cer species were increased (Tukey’s HSD, p < 0.05) (**Fig 4A-D**), whilst GSLs (HexCers, GlcCers) were decreased (Tukey’s HSD, p < 0.05) in the ‘advanced’ fibrosis (NASH F3) as compared to ‘minimal’ and / or ‘mild’ disease stages (**Fig 4E-F**). Overall, the directional changes (up or downregulated) of the predicted RMs of GSLs in the liver were comparable with the differences (increase or decrease) in the serum GSL levels of the patients at different stages of NASH-associated fibrosis. Intriguingly, there was an abrupt decrease in the levels of Cers and HexCers at NASH F4 (cirrhosis) stage, which may be attributed to the alterations in the SL pathway(s) in the liver.

Serum levels of retinol, retinol palmitate and cholecalciferol (D3) across various stages of NAFLD are shown in (**Fig S10**).

### Impact of genetic variants associated with NAFLD on liver metabolic pathways

Genetic susceptibility plays a vital role in the development of NAFLD (Anstee et al., 2020; Trepo and Valenti, 2020). Here, we studied three of the most widely validated NAFLD-associated common genetic variants, *i*.*e*., PNPLA3 (rs738409), TM6SF2 (rs58542926) and HSD17B13 (rs72613567) (Anstee et al., 2020; Liu et al., 2014; Luukkonen et al., 2017; Ma et al., 2019; Trepo and Valenti, 2020). Some of these gene variants are known to alter TG hydrolysis (He et al., 2010) and retinol (Vitamin A) metabolism (Kovarova et al., 2015; Mondul et al., 2015) and increase the severity of NAFLD (He et al., 2010; Luukkonen et al., 2017; Pettinelli et al., 2018). However, little is known about the metabolic differences conferred by these variants.

We developed personalized GEMs for hepatocytes in the individuals carrying one of these three common gene variants, *PNPLA3* (GC, GG) (n=69), *TM6SF2* (CT, TT) (n=13), *HSD17B13* (-T, TT) (n=21) compared with wild type (WTs) (n=36), *i*.*e*., individuals homozygous for all the three gene variants: *PNPLA3 (CC), TM6SF2 (CC) and HSD17B13 (--)* (**Tables 1** and **Fig S11**), using genome-wide transcriptomics data and the *iHepatocytes2322* model. Selection of individuals exclusively carrying *PNPLA3, TM6SF2* or *HSD17B13* gene variants from the study cohort (Govaere et al., 2020) is given in (**Table S1**). The occurrence scores of the metabolic tasks carried out by the gene variants and wild type (WT) models are shown in (**Fig S12**).

Pairwise PLS-DA analysis of the fluxes across the metabolic subsystems predicted by the hepatocyte-GEMs of *PNPLA3, TM6SF2* or *HSD17B13* variant carriers *vs*. WT showed remarkable separation between these groups, with AUCs = [0.86, 0.85 and 0.84], respectively (**Fig 6A-C**). PLS-DA identified five metabolic subsystems, fatty acid biosynthesis and oxidation, oxidative phosphorylation, terpinoid biosynthesis and folate metabolism, which were commonly altered (VIP > 0.1) (**Fig 6D** and **S13**) between the gene variants and the WT. In addition, a pairwise comparision between the PNPLA3 variants and WT groups suggest that GSL metabolism are downregulated (two-sample t-test, p<0.05 corrected for FDR) whilst cysteine and methionine, and vitamin B6 metabolism was upregulated (p<0.05 corrected for FDR). *PNPLA3* and *HSD17B13* variants showed a decrease (p<0.05 corrected for FDR) in the β-oxidation of fatty acids as compared to the WT, whilst *TM6SF2* variant showed an elevation in the cholesterol biosynthesis (**Fig 6E)**.

**Figure 6.**
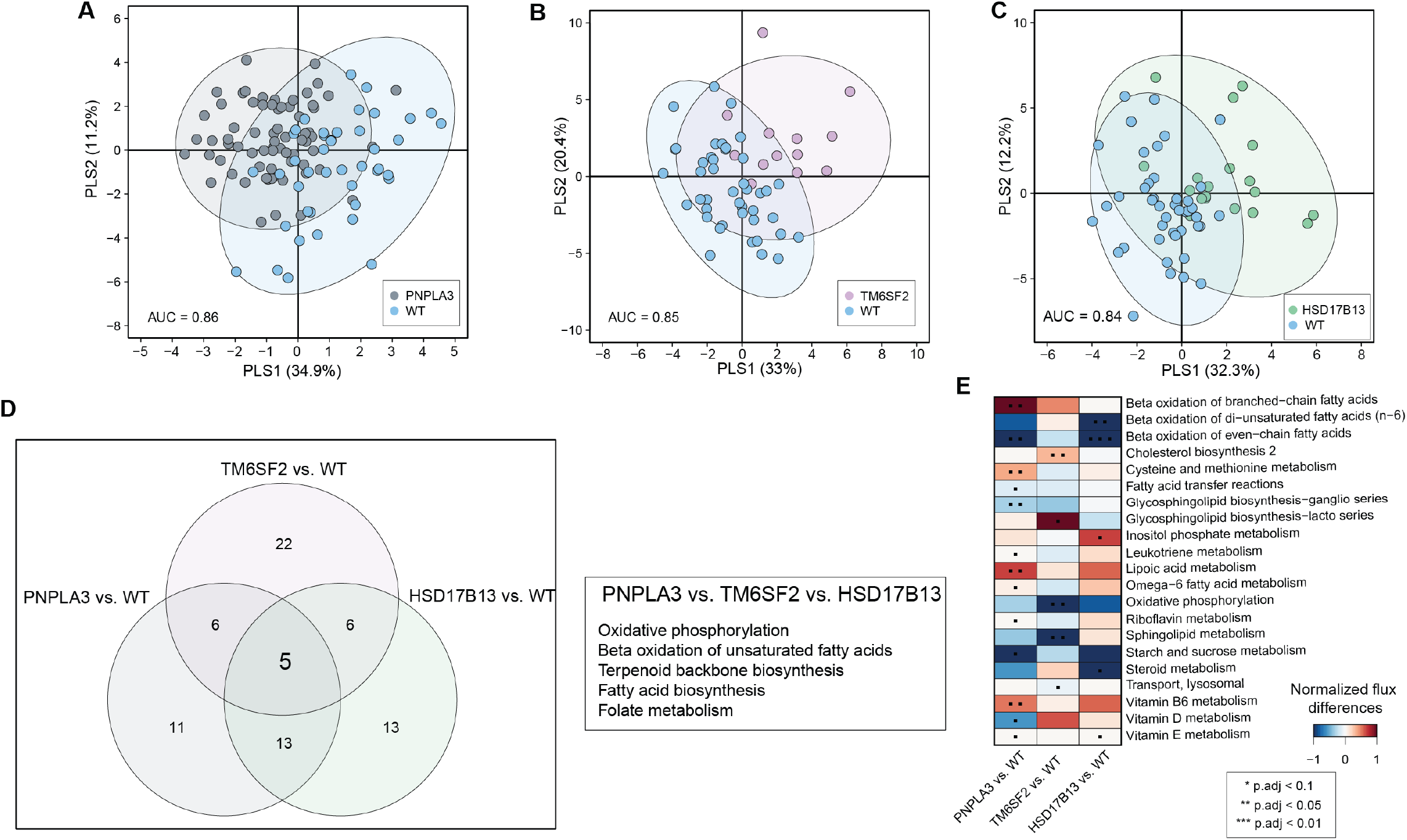
Metabolic pathway differences across the genetic modifiers of NAFLD. PLS-DA–score plot showing metabolic pathway differences across three major gene variants vs. WT groups. (**A**) *PNPLA3* vs. WT. (**B**) *TM6SF2* vs. WT. (**C**) *HSD17B13* vs. WT. (**D**) Metabolic pathways/subsystems commonly regulated among three different gene variants with (PLS-DA, VIP score >1). (**E**) Normalized flux differences across the metabolic subsystems between three major gene variants associated vs. WT groups. Red and blue color denotes up- and downregulation (two-sample t-test, p<0.05 corrected for FDR) of reaction fluxes across the metabolic subsystems within at two different stages of NAFLD.

## Materials and Methods

### Subject Details

A full description of the transcriptomic analysis of the whole liver RNA-Seq data used in this study has previously been reported (Govaere et al., 2020). Patient samples were derived from the European NAFLD Registry (NCT04442334) (Hardy et al., 2020) and comprised snap-frozen biopsy samples and associated clinical data from 206 patients diagnosed with histologically characterized NAFLD in France, Germany, Italy, or the United Kingdom. All biopsies samples obtained in this study were centrally scored by two expert liver pathologists according to the semiquantitative NASH-Clinical Research Network ‘NAFLD Activity Score’ (NAS) (Kleiner et al., 2005). Fibrosis was staged from F0 to F4 (cirrhosis) (Govaere et al., 2020). Patients with alternate diagnoses and etiologies were excluded, including excessive alcohol intake (30 g per day for males and 20 g per day for females), viral hepatitis, autoimmune liver diseases, and steatogenic medication use. As previously described (Hardy et al., 2020), collection and use of samples and clinical data for this study were approved by the relevant local and/or national Ethical Review Committee covering each participating center, with all patients providing informed consent for participation. All participant recruitment and informed consent processes at recruitment centers were conducted in compliance with nationally accepted practice in the respective territory and in accordance with the World Medical Association Declaration of Helsinki 2018.

### Transcriptomics

Transcriptomic (RNA-seq) data associated with this study was obtained from (Govaere et al., 2020); also available in the NCBI GEO repository (accession GSE135251). Differentially expressed genes between a paired conditions were either obtained (Govaere et al., 2020), or estimated by a method stated in (Govaere et al., 2020). All genes that were differentially expressed between the case vs. control at (p-values corrected for FDR < 0.05) were included in the RM analysis.

### Genotyping

Genotypes were obtained from the GWAS data described in detail in (Anstee et al., 2020). Some data was also obtained from RNA-seq as reported elsewhere (Govaere et al., 2020).

### Analysis of lipids and polar metabolites

#### Lipidomics analysis

Serum samples were randomized and extracted using a modified version of the previously-published Folch procedure (Nygren et al., 2011). In short, 10 µL of 0.9% NaCl and, 120 µL of CHCl3: MeOH (2:1, v/v) containing the internal standards (c = 2.5 µg/mL) was added to 10 µL of each serum sample. The standard solution contained the following compounds: 1,2-diheptadecanoyl-sn-glycero-3-phosphoethanolamine (PE(17:0/17:0)), N-heptadecanoyl-D-erythro-sphingosylphosphorylcholine (SM(d18:1/17:0)), N-heptadecanoyl-D-erythro-sphingosine (Cer(d18:1/17:0)), 1,2-diheptadecanoyl-sn-glycero-3-phosphocholine (PC(17:0/17:0)), 1-heptadecanoyl-2-hydroxy-sn-glycero-3-phosphocholine (LPC(17:0)) and 1-palmitoyl-d31-2-oleoyl-sn-glycero-3-phosphocholine (PC(16:0/d31/18:1)), were purchased from Avanti Polar Lipids, Inc. (Alabaster, AL, USA), and, triheptadecanoylglycerol (TG(17:0/17:0/17:0)) was purchased from Larodan AB (Solna, Sweden). The samples were vortex mixed and incubated on ice for 30 min after which they were centrifuged (9400 × g, 3 min). 60 µL from the lower layer of each sample was then transferred to a glass vial with an insert and 60 µL of CHCl3: MeOH (2:1, v/v) was added to each sample. The samples were stored at −80 °C until analysis.

Calibration curves using 1-hexadecyl-2-(9Z-octadecenoyl)-sn-glycero-3-phosphocholine (PC(16:0e/18:1(9Z))), 1-(1Z-octadecenyl)-2-(9Z-octadecenoyl)-sn-glycero-3-phosphocholine (PC(18:0p/18:1(9Z))), 1-stearoyl-2-hydroxy-sn-glycero-3-phosphocholine (LPC(18:0)), 1-oleoyl-2-hydroxy-sn-glycero-3-phosphocholine (LPC(18:1)), 1-palmitoyl-2-oleoyl-sn-glycero-3-phosphoethanolamine (PE(16:0/18:1)), 1-(1Z-octadecenyl)-2-docosahexaenoyl-sn-glycero-3-phosphocholine (PC(18:0p/22:6)) and 1-stearoyl-2-linoleoyl-sn-glycerol (DG(18:0/18:2)), 1-(9Z-octadecenoyl)-sn-glycero-3-phosphoethanolamine (LPE(18:1)), N-(9Z-octadecenoyl)-sphinganine (Cer(d18:0/18:1(9Z))), 1-hexadecyl-2-(9Z-octadecenoyl)-sn-glycero-3-phosphoethanolamine (PE(16:0/18:1)) from Avanti Polar Lipids, 1-Palmitoyl-2-Hydroxy-sn-Glycero-3-Phosphatidylcholine (LPC(16:0)), 1,2,3 trihexadecanoalglycerol (TG(16:0/16:0/16:0)), 1,2,3-trioctadecanoylglycerol (TG(18:0/18:0/18:)) and 3β-hydroxy-5-cholestene-3-stearate (ChoE(18:0)), 3β-Hydroxy-5-cholestene-3-linoleate (ChoE(18:2)) from Larodan, were prepared to the following concentration levels: 100, 500, 1000, 1500, 2000 and 2500 ng/mL (in CHCl3:MeOH, 2:1, v/v) including 1250 ng/mL of each internal standard.

The samples were analyzed by ultra-high-performance liquid chromatography quadrupole time-of-flight mass spectrometry (UHPLC-QTOFMS). Briefly, the UHPLC system used in this work was a 1290 Infinity II system from Agilent Technologies (Santa Clara, CA, USA). The system was equipped with a multi sampler (maintained at 10 °C), a quaternary solvent manager and a column thermostat (maintained at 50 °C). Injection volume was 1 µL and the separations were performed on an ACQUITY UPLC® BEH C18 column (2.1 mm × 100 mm, particle size 1.7 µm) by Waters (Milford, MA, USA). The mass spectrometer coupled to the UHPLC was a 6545 QTOF from Agilent Technologies interfaced with a dual jet stream electrospray (Ddual ESI) ion source. All analyses were performed in positive ion mode and MassHunter B.06.01 (Agilent Technologies) was used for all data acquisition. Quality control was performed throughout the dataset by including blanks, pure standard samples, extracted standard samples and control serum samples, including in-house serum and a pooled QC with an aliquot of each sample was collected and pooled and used as quality control sample.

Relative standard deviations (% RSDs) for identified lipids in the control serum samples (n = 13) and in the pooled serum samples (n = 54) were on average 22.4% and 17.5%, respectively.

Mass spectrometry data processing was performed using the open source software package MZmine 2.18 (Pluskal et al., 2010). The following steps were applied in this processing: (i) Crop filtering with a m/z range of 350 – 1200 m/z and an RT range of 2.0 to 12 minutes, (ii) Mass detection with a noise level of 750, (iii) Chromatogram builder with a minimum time span of 0.08 min, minimum height of 1000 and a m/z tolerance of 0.006 m/z or 10.0 ppm, (iv) Chromatogram deconvolution using the local minimum search algorithm with a 70% chromatographic threshold, 0.05 min minimum RT range, 5% minimum relative height, 1200 minimum absolute height, a minimum ration of peak top/edge of 1.2 and a peak duration range of 0.08 - 5.0, (v), Isotopic peak grouper with a m/z tolerance of 5.0 ppm, RT tolerance of 0.05 min, maximum charge of 2 and with the most intense isotope set as the representative isotope, (vi) Peak filter with minimum 12 data points, a FWHM between 0.0 and 0.2, tailing factor between 0.45 and 2.22 and asymmetry factor between 0.40 and 2.50, (vii) Join aligner with a m/z tolerance of 0.009 or 10.0 ppm and a weight for of 2, a RT tolerance of 0.1 min and a weight of 1 and with no requirement of charge state or ID and no comparison of isotope pattern, (viii) Peak list row filter with a minimum of 10% of the samples (ix) Gap filling using the same RT and m/z range gap filler algorithm with an m/z tolerance of 0.009 m/z or 11.0 ppm, (x) Identification of lipids using a custom database search with an m/z tolerance of 0.009 m/z or 10.0 ppm and a RT tolerance of 0.1 min, and (xi) Normalization using internal standards PE(17:0/17:0), SM(d18:1/17:0), Cer(d18:1/17:0), LPC(17:0), TG(17:0/17:0/17:0) and PC(16:0/d30/18:1)) for identified lipids and closest ISTD for the unknown lipids followed by calculation of the concentrations based on lipid-class concentration curves. Identification of lipids was based on in house laboratory based on LC-MS/MS data on retention time and mass spectra.

#### Analysis of polar metabolites

Serum samples were randomized and sample preparation was carried out as described previously (Castillo et al., 2011). The maternal samples were analysed as one batch and the cord blood samples as a second batch. In summary, 400 μL of MeOH containing ISTDs (heptadecanoic acid, deuterium-labeled DL-valine, deuterium-labeled succinic acid, and deuterium-labeled glutamic acid, c = 1 µg/mL) was added to 30 µl of the serum samples which were vortex mixed and incubated on ice for 30 min after which they were centrifuged (9400 × g, 3 min) and 350 μL of the supernatant was collected after centrifugation. The solvent was evaporated to dryness and 25 μL of MOX reagent was added and the sample was incubated for 60 min at 45 °C. 25 μL of MSTFA was added and after 60 min incubation at 45 °C 25 μL of the retention index standard mixture (n-alkanes, c=10 µg/mL) was added.

The analyses were carried out on an Agilent 7890B GC coupled to 7200 QTOF MS. Injection volume was 1 µL with 100:1 cold solvent split on PTV at 70 °C, heating to 300 °C at 120 °C/minute. Column: Zebron ZB-SemiVolatiles. Length: 20m, I.D. 0.18mm, film thickness: 0.18 µm. With initial Helium flow 1.2 mL/min, increasing to 2.4 mL/min after 16 mins. Oven temperature program: 50 °C (5 min), then to 270°C at 20 °C/min and then to 300 °C at 40 °C/min (5 min). EI source: 250 °C, 70 eV electron energy, 35µA emission, solvent delay 3 min. Mass range 55 to 650 amu, acquisition rate 5 spectra/s, acquisition time 200 ms/spectrum. Quad at 150 °C, 1.5 mL/min N2 collision flow, aux-2 temperature: 280 °C.

Calibration curves were constructed using alanine, citric acid, fumaric acid, glutamic acid, glycine, lactic acid, malic acid, 2-hydroxybutyric acid, 3-hydroxybutyric acid, linoleic acid, oleic acid, palmitic acid, stearic acid, cholesterol, fructose, glutamine, indole-3-propionic acid, isoleucine, leucine, proline, succinic acid, valine, asparagine, aspartic acid, arachidonic acid, glycerol-3-phosphate, lysine, methionine, ornithine, phenylalanine, serine and threonine purchased from Sigma-Aldrich (St. Louis, MO, USA) at concentration range of 0.1 to 80 μg/mL. An aliquot of each sample was collected and pooled and used as quality control samples, together with a NIST SRM 1950 serum sample and an in-house pooled serum sample. Relative standard deviations (% RSDs) of the metabolite concentrations in control serum samples showed % RSDs within accepted analytical limits at averages of 27.2% and 29.2% for in-house QC abd pooled QC samples.

### Genome-scale metabolic modeling

#### Development of stage-specific GEMs of human hepatocytes along the NAFLD spectrum

Stage-specific functional GEMs of human hepatocytes were developed by step-wise combining *iMAT (Zur et al*., *2010)* and *E-Flux* (Colijn et al., 2009) algorithms, applied to *iHepatocytes2322* as a template model (GEM). *iMAT* approach finds the optimal trade-off between inclusion and exclusion of high and low-expression reactions. It does not explicitly depends on an metabolic objective function (Opdam et al., 2017; Zur et al., 2010). *iMAT* requires three sets of input reactions such as high, low and moderately expressed. The reaction weights were determined by integrating the NAFLD stage-specific gene expression data to *iHepatocytes2322* using gene-protein-reaction associations (GPR) rules. To categorize the model reactions into high, low and moderately expressed, we estimated the (mean ± sd) of the log-normal distribution of the reaction weights / expression (Opdam et al., 2017; Zur et al., 2010). Accordingly, the feasibility of a particular reaction(s) to be included or discarded in the draft model was determined. Mixed-integer linear programming (MILP) was used to determine the functionality of a model, *i*.*e*., reactions that can carry fluxes were kept and blocked reactions were rectified or removed. The *‘FASTCC’* algorithm implemented in COnstraint-Based Reconstruction and Analysis Toolbox (COBRA Toolbox v3.0)(Heirendt et al., 2017) was used for testing the functionality / consistency of the model. Next, *E-Flux* (Colijn et al., 2009) algorithm was used to apply constrain to these models by using transcriptomics data. *E-Flux* depends on a metabolic objective function, maximization of lipid droplet accumulation via ‘*reaction ID: HMR_0031’* in *iHepatocytes2322* model was set as an objective function. All the models were tested to carry out 256 metabolic tasks as given in (Mardinoglu et al., 2013; Mardinoglu et al., 2014), using *‘CheckTask’* function implemented in the Reconstruction, Analysis and Visualization of Metabolic Networks) (RAVEN) 2.0 suite (Wang et al., 2018). The draft models were manually curated.

Mixed integer linear programming (MILP) was performed using ‘MOSEK 8’ solver (licensed for the academic user) integrated in the RAVEN 2.0 suite (Wang et al., 2018). Linear programming (LP) and optimization was performed using *‘ILOG-IBM CPLEX (version 128)’* solver. Quality control and sanity checks (Thiele and Palsson, 2010) were performed using Cobra toolbox v3.0 (Heirendt et al., 2017). Simulations were performed using Cobra toolbox v3.0 (Heirendt et al., 2017) and RAVEN 2.0 suite (Wang et al., 2018). All the operations were performed in MATLAB 2017b (Mathworks, Inc., Natick, MA, USA).

By integrating NAFLD stage-specific whole tissue transcriptomics (RNASeq) data with *iHepatocytes2322*, we contextualized and developed hepatocyte-GEMs for different stages of NASH-associated fibrosis, *i*.*e*., ‘minimal’ NAFL + NASH (F0-1) (n= 85); ‘mild’ NAFL + NASH (F0-2) (n= 138); ‘clinically significant’ non-cirrhotic fibrosis (NASH F2-3) (n= 107); and ‘advanced’ fibrosis (NASH F3-4) (n= 68). A similar approach was adapted to contextualize GEMs (*iHepatocytes2322*) for hepatocytes in the individuals exhibiting carriage of gene variants, *PNPLA3* (GC, GG) (n=69), *TM6SF2* (CT, TT) (n=13), *HSD17B13* (-T, TT) (n=21) and wild type (WTs) (n=36) (**Table 1** and **S1**).

#### Normalized flux differences

We deployed an unbiased and non-uniform random sampling (RS) method (Bordel et al., 2010) that finds solutions among the feasible flux distributions of the metabolic network. RS does not explicitly depends on an metabolic objective function (Bordel et al., 2010; Herrmann et al., 2019). By applying RS on the context / stage-specific GEMs, we estimated the flux distribution (sampled, n=1000). RS was performed using RAVEN 2.0 suite using *‘MOSEK 8’* solver (Wang et al., 2018). Only reactions having non-zero measurable fluxes (>1e-6 mmol/gDW/hr) fluxes were considered in the pairwise (case – control) comparision. The absolute values of the reaction fluxes were grouped by their corresponding subsystems or pathways. The fluxes per subsystem were averaged and subjected to quantile normalization. Thereby, the flux differences between the case *vs*. control was estimated by performing a two sample t-test applied to log-normal distribution of data. The p-values were subjected to multiple testing and False Discovery Rates (FDR) corrections. FDR corrected p-value < 0.05 was considered as statistically significant.

#### Partial least squares discriminant analysis

Multiclass and two groups partial least squares discriminant analysis (PLS-DA) (Le Cao et al., 2011) models were developed across the NAFLD spectrum, and for the three major gene variants respectively; by using the ‘*plsda’* function coded in the *‘mixOmics v6*.*3*.*2’* package, implemented in R statistical programming language (R Development Core Team, 2018). Variable Importance in Projection (VIP) scores (Farrés et al., 2015) of the features (subsystems) were estimated. PLS-DA models were cross-validated (Westerhuis et al., 2008) by 7-fold cross-validation and models diagnostics were generated using *‘perf’* and *‘auroc’* functions. All features that passed (VIP scores > 1) were listed as significantly altered between or among the groups.

#### Reporter metabolite analysis

RM analysis (Cakir et al., 2006; Patil and Nielsen, 2005) was performed by using ‘*reporterMetabolites’* function of the RAVEN 2.0 suite (Wang et al., 2018). As the hepatocyte GEMs were parameterized by the transcriptomics data; determined by the two-step contextualization approach, the boundaries of the exchange reactions were not relaxed between [-inf, +inf]. Multiple testing was performed and the nominal p-values were subjected to FDR corrections. The RMs that passed the threshold (FDR corrected p-values < 0.05) were considered as significantly altered between the differential condition (case – control).

#### Analysis of metaboliomics data

The metabolomics dataset was log_2_ transformed. Homogeneity of the samples were assessed by principal component analysis (PCA) (Carey et al., 1975) and no outliers were detected (95% confidence interval). The log-normalized intensities of the total identified lipids and polar metabolites were stratified into various NAFLD groups (**Table S1**). Multiple comparision was performed using one-way analysis of variance (ANOVA), followed by Tukey’s honest significant difference (HSD) – a post hoc test. p-values (HSD) < 0.05 was considered as statistically significant.

#### Data visualization and graphics

Several libraries / packages of R v3.6.0, a statistical programming language such as *‘Heatmap*.*2’, ‘boxplot’, ‘beanplot’*, ‘*gplot*’, and ‘*ggplot2*’ and *Cytoscape v3*.*8*.*0 software* were used for data visualization and network analysis.

## Discussion

GSMM of human hepatocytes in NASH-associated fibrosis (n=206) identified several metabolic signatures and pathways markedly regulated at different stages of fibrosis. The previous GSMM study of human NAFLD (Mardinoglu et al., 2014) also identified altered metabolic pathways in NAFLD, however, that study was confined to a relatively small patients group, not representing the full spectrum of NAFLD. Here, we used genome-wide transcriptomics (RNA-seq) (Govaere et al., 2020) data that provided a dynamic range of genes, enzymes / reactions, metabolites and their interactions, to model stage-specific GEMs for human hepatocytes in NAFLD (**Fig 1**). In addition, the study cohort allowed us to stratify and model the metabolic differences in three major genetic variants (*PNPLA3, TM6SF2* and *HSD17B13*) associated with risk and severity of NAFLD.

Our results implicate changes in the levels of vitamins (A, E) in the NASH-associated fibrosis progression. In the hepatocytes, a significant increase in the RMs of retinoic acid derivatives were observed in NASH F2, F3 (*vs*. NAFL) and in ‘clinically significant’ non-cirrhotic fibrosis (*vs*. ‘minimal disease’). A small increase in the serum levels of Vitamin A (retinol) and retinyl palmitate was observed at NASH F0-1 stage, whilst retinyl palmitate was markedly decreased at NASH F4 stage. At the F4 stage, the activated hepatic stellate cells tend to lose the retinyl esters stored in the liver, ultimately leading to vitamin A deficiency (Pettinelli et al., 2018; Saeed et al., 2017). Vitamin A is a regulator of glucose and lipid metabolism in the liver and adipose tissue and may attenuate in the development of NAFLD (Pettinelli et al., 2018; Saeed et al., 2017). The PNPLA3 gene product is known to have retinyl ester hydrolase activity, and *PNPLA3-I148M* is associated with low serum retinol level with enhanced retinyl esters in the liver of NAFLD patients (Kovarova et al., 2015; Mondul et al., 2015). However, GSMM has not identified any differences in vitamin A metabolism related to *PNPLA3* variant carriage. Intriguingly, Vitamin E derivatives were subsequently decreased across the stages of fibrosis. High dose of Vitamin E supplementation has been shown to improve histological steatohepatitis over placebo in the PIVENS randomized controlled trial of pioglitazone or vitamin E in patients with NASH (Sanyal et al., 2010).

We observed a dynamic regulation of complex GSLs in the advanced fibrosis. At that stage, the majority of cerebrosides (HexCers: glucosylceramides (GlcCers) and lactosylceramides (LacCers)), and globosides were decreased in the patient groups. Intriguingly, the serum concentrations of GSLs (HexCers, GlcCers) were markedly decreased in the patients with advanced fibrosis or cirrhosis (NASH F4). Cers are the key intermediates of sphingolipid metabolism that promote cellular proliferation, differentiation and cell death (Gault et al., 2010; Turpin-Nolan and Bruning, 2020). Cers interact with several pathways involved in insulin resistance, oxidative stress, inflammation, and apoptosis, that are linked to NAFLD (Gault et al., 2010; Pagadala et al., 2012). Understanding the role of Cers in the staging of NASH-associated fibrosis is of great diagnostic interest (Pagadala et al., 2012). In normal physiological conditions, Cers converts to GSLs, which prevents its excessive accumulation in the cells (Ishay et al., 2020). Our data suggest that in patients with advanced fibrosis, the serum concentrations of Cers increase up to NASH F3, and decrease at NASH F4 stages. Differential flux analysis showed an increase of Cer production via *de novo* pathway, whilst production of GSLs (GlcCer, GalCer and LacCers) decreased in patients with advanced fibrosis. Thus, the conversion of Cers to GSLs (*via* glucosylceramide synthase, GCS) might have been compromised in ‘advanced’ fibrosis. Intriguingly, we found that the GSLs (LacCers and globosides) were associated with GAGs (heparin, keratin sulphate), which was also altered in the progression of NASH-associated fibrosis. It is well demonstrated that inhibiting GSL synthesis in obese mice has improved glucose homeostasis and markedly reduced the development of NAFL (Zhao et al., 2009).

Taken together, we identified several novel metabolic signatures and pathways in progressive stages of NAFLD, which enabled us to understand how different metabolites and their intermediates are regulated across various stages of NASH-associated fibrosis. While some of our key results corroborate with the earlier findings, others allowed us to elucidate the dys(regulation) of metabolic pathways in NAFLD. Moreover, GSMM has demonstrated several stage-specific metabolic changes, which might help discover biomarkers, identify drug targets, and ultimately lead to effective therapeutic strategies for NASH. To this end, our data indicate the significance of GSL pathways and targeting these pathways might ameliorate the liver pathology associated with NAFL and NASH-associated fibrosis. However, some of these findings remains to be validated by *in vivo* and/or *in vitro* experiments.

## Data Availability

Scripts for GSMM, contextualization and data analysis can be downloaded from (https://github.com/parthoBTK/Personalized_Liver_Models.git). The personalized GEMs (.mat files) of human-hepatocytes contextualized for different stages of NAFLD, and three major gene variants are available upon request. The RNA-Seq data are available in the NCBI GEO repository (accession GSE135251). The lipidomic / metabolomic datasets generated in this study were submitted to the Metabolomics Workbench repository (https://www.metabolomicsworkbench.org) and were assigned a project ID PR001091 (doi: 10.21228/M8P97V).

## Acknowledgements

We would like to thank the EPoS (Elucidating Pathways of Steatohepatitis) (epos-nafld.eu) investigators.

This study has been supported by the EPoS (Elucidating Pathways of Steatohepatitis) consortium funded by the Horizon 2020 Framework Program of the European Union under Grant Agreement 634413, the LITMUS (Liver Investigation: Testing Marker Utility in Steatohepatitis) consortium funded by the Innovative Medicines Initiative (IMI2) Program of the European Union under Grant Agreement 777377. Q.M.A. is a Newcastle NIHR Biomedical Research Centre investigator. T.H. and M.O. received funding from the Novo Nordisk Foundation (Grant no. NNF20OC0063971). M.O. was also supported by the Academy of Finland (grant no. 333981). T.V.P. laboratory was funded by the MRC MDU programme grant.

## Author Contributions

M.O. proposed and design the study. Q.M.A., A.K.D and M.O. assisted with the formulation of the study design. M.O., Q.M.A., A.K.D., and T.H. supervised the study. P.S. performed GSMM and statistical analysis. T.S. and D.G. performed metabolomics analysis, supervised by T.H. A.M. performed metabolomics data analysis. O.G. and S.C. performed the transcriptomic analysis, supervised by A.K.D. V.R. E.B. J.M.S., A.V.-P., M.A., Q.M.A. contributed to clinical data acquisition in the EPoS cohort, coordinated by Q.M.A.. P.S. and M.O. wrote the manuscript. All authors critically revised the manuscript for intellectual content and approved the final manuscript.

M.O. is the guarantor of this work and, as such, had full access to all of the data in the study and takes responsibility for the integrity of the data and the accuracy of the data analysis.

## Supplemental Information

Supplemental Information includes 13 figures (**Fig S1-S13**) and one table (**Table S1**).

## Supplemental Material

**Figure S1.**
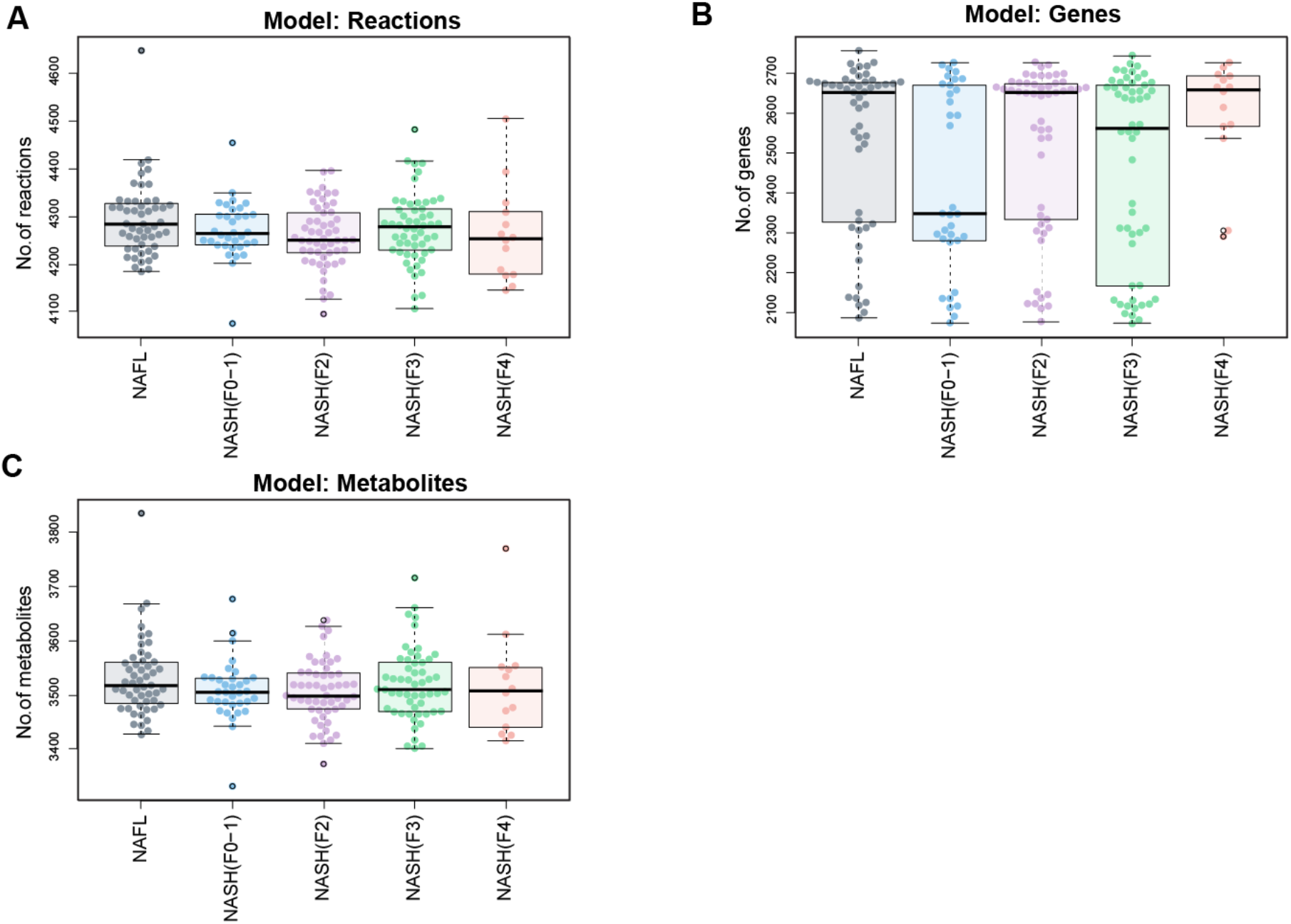
Personalized genome-scale metabolic models of human hepatocytes in NAFLD. Boxplots showing distribution of number of reactions (**A**), genes (**B**) and metabolites (**C**) (marked by dots) contained in the genome-scale metabolic models (GEMs) of human hepatocytes developed for (n=206) subjects and contextualized for various stages of NAFLD using transcriptomics data.

**Figure S2.**
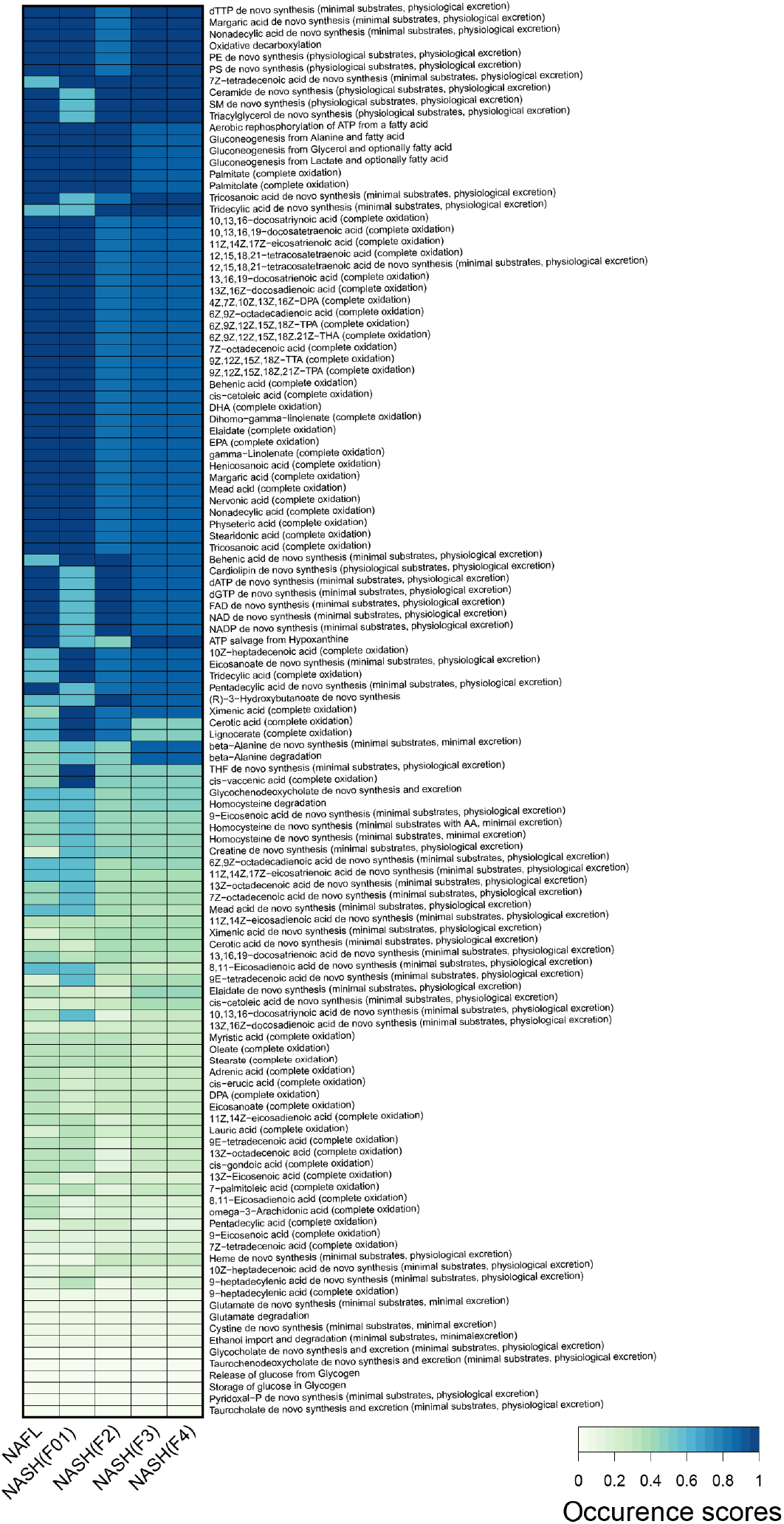
Metabolic tasks carried out by the NAFLD stage-specific personalized GEMs. [0-1] denotes low to high occurrence scores (OS). OS acquired by a GEM is based on its capability (‘Yes’ or ‘No’) to perform the assigned metabolic task.

**Figure S3.**
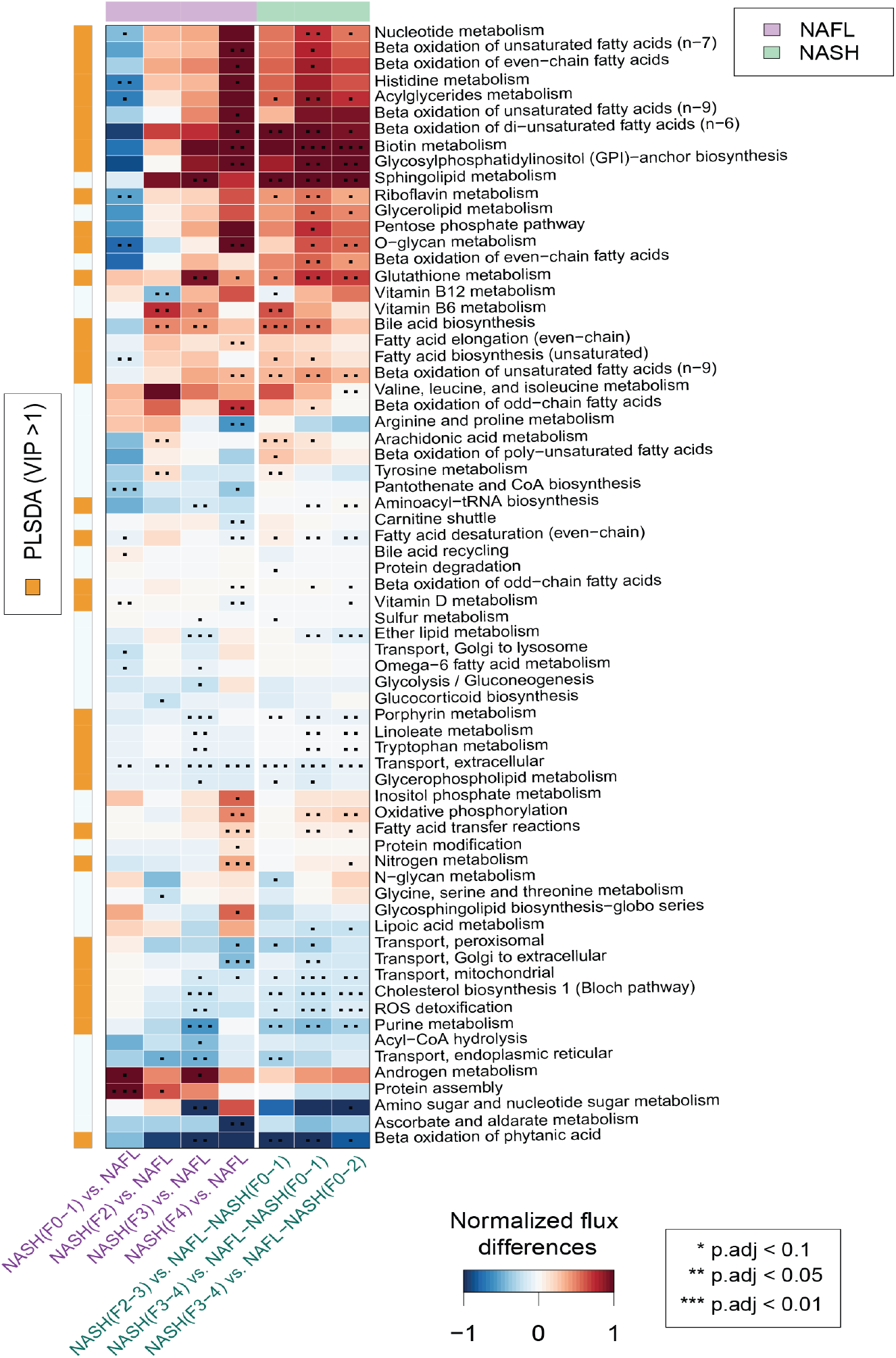
Predicted flux differences across the metabolic subsystems of human hepatocytes at different stages of NAFLD. Red and blue colors denotes up- and downregulated fluxes across the metabolic subsystems within two different stages of NAFLD. ‘**’ and ‘***’ denotes statistical signifance (two-sample t-test, p.adj<0.05 and p.adj<0.01 corrected for FDR) respectively. White color denotes ‘no change’. Flux state at each stage was estimated by random sampling.

**Figure S4.**
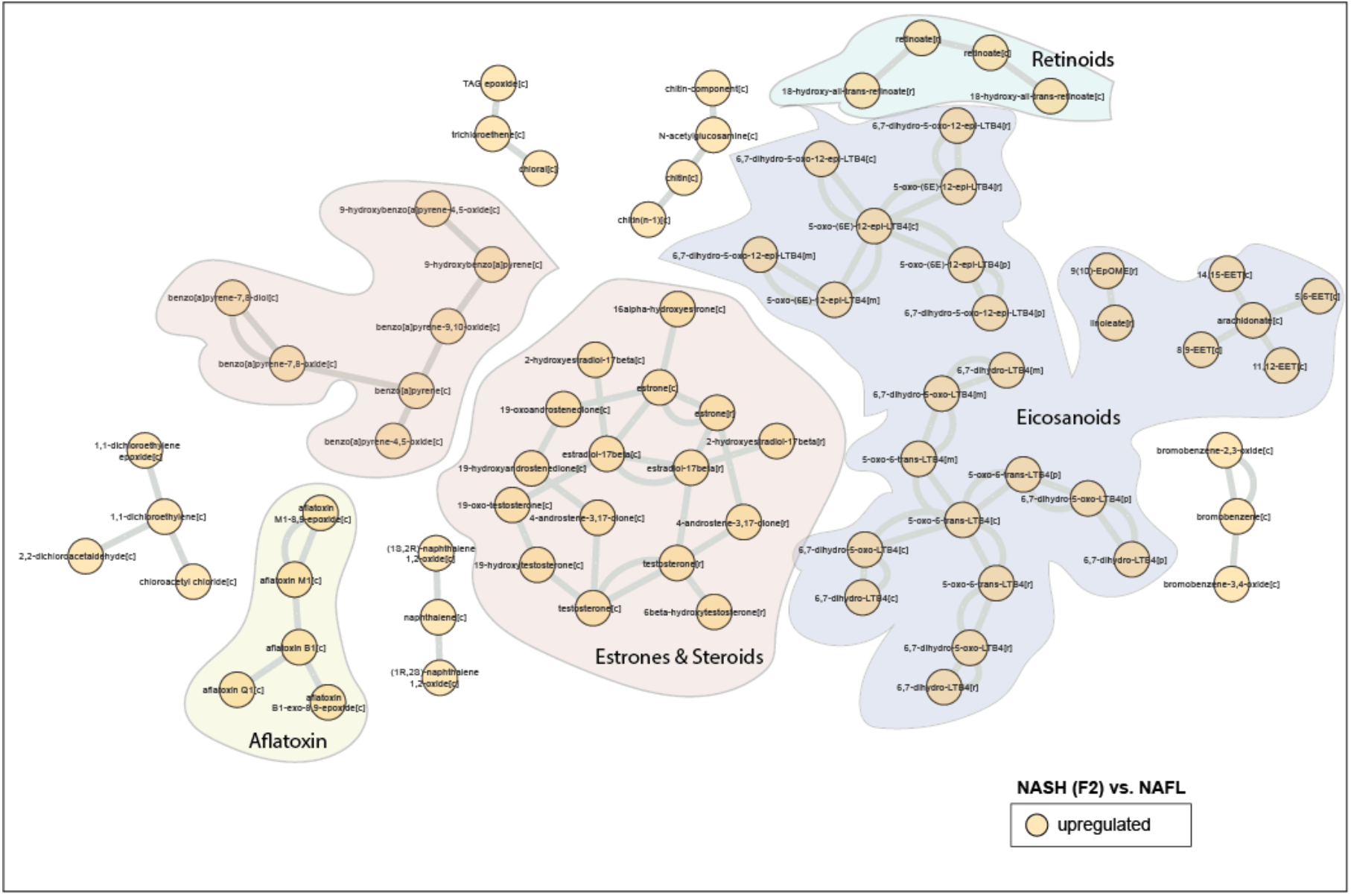
A metabolic-centric view of reporter metabolite (RM) modules that were significantly altered between NASH F2 *vs*. NAFL. Orange color denotes upregulated (p<0.05 corrected for FDR). Each node represents a ‘RM’ and single or double lines represent reversible or irreversible metabolic reactions respectively. RMs that belong to a particular chemical class are color coded.

**Figure S5.**
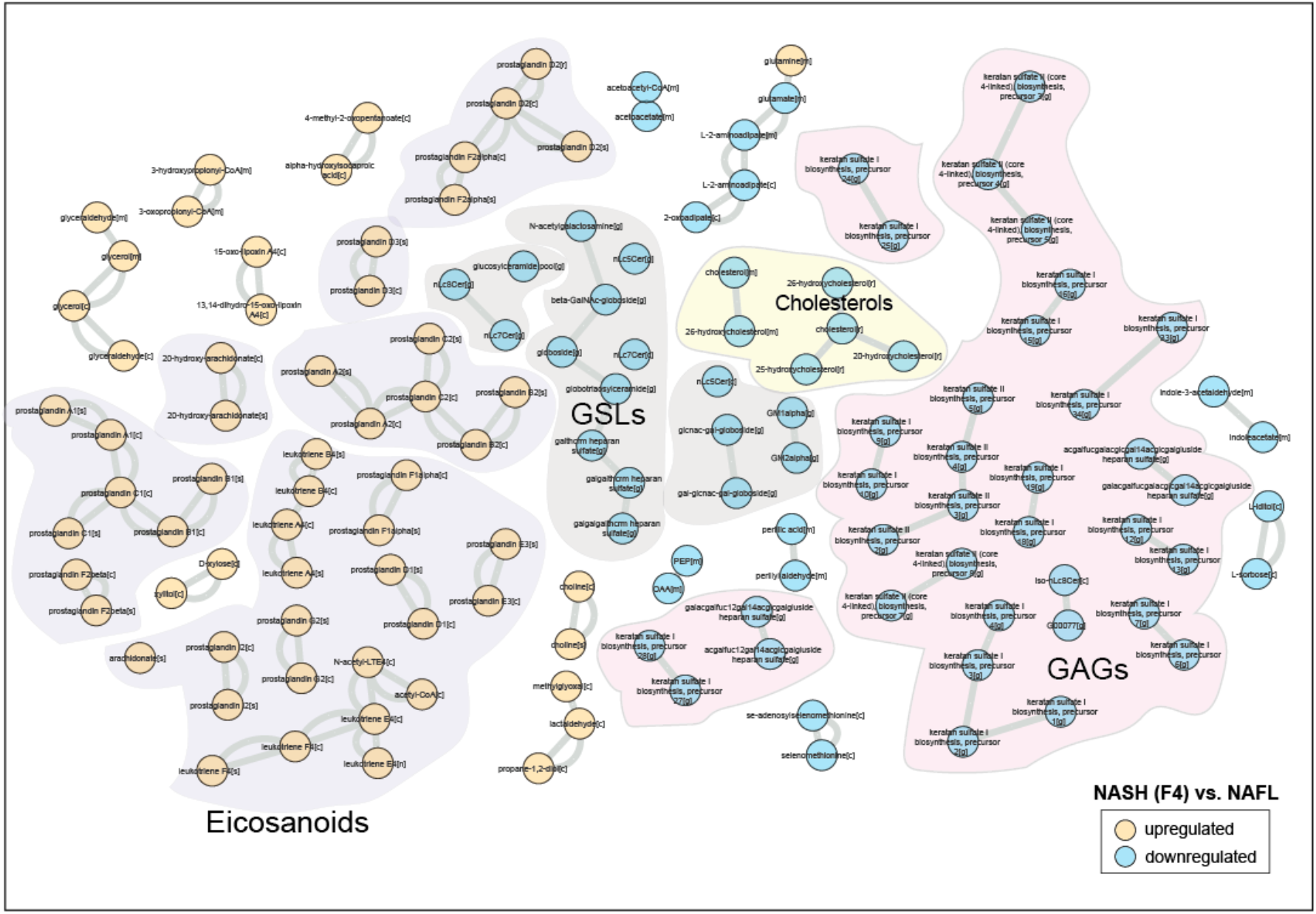
A metabolic-centric view of reporter metabolite (RM) modules that were significantly altered between NASH F4 *vs*. NAFL. Orange and blue color denotes up- and downregulated (p<0.05 corrected for FDR) respectively. Each node represents a ‘RM’ and single or double lines represent reversible or irreversible metabolic reactions respectively. RMs that belong to a particular chemical class are color coded.

**Figure S6.**
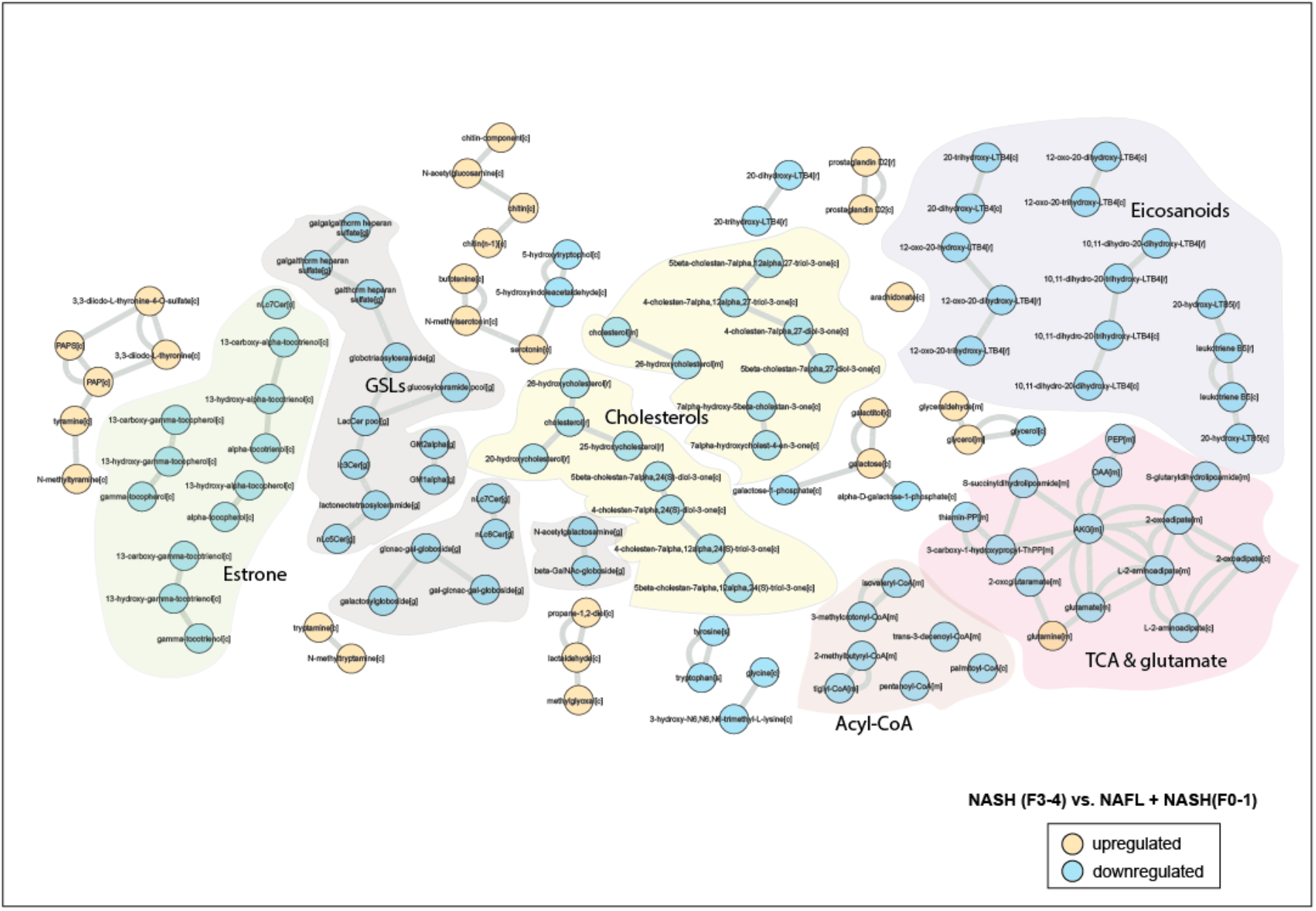
A metabolic-centric view of reporter metabolite (RM) modules that were significantly altered between ‘advanced’ fibrosis *vs*. ‘minimum’ disease. Orange and blue color denotes up and downregulated (p<0.05 corrected for FDR) respectively. Each node represents a ‘RM’ and single or double lines represent reversible or irreversible metabolic reactions respectively. RMs that belong to a particular chemical class are color coded.

**Figure S7.**
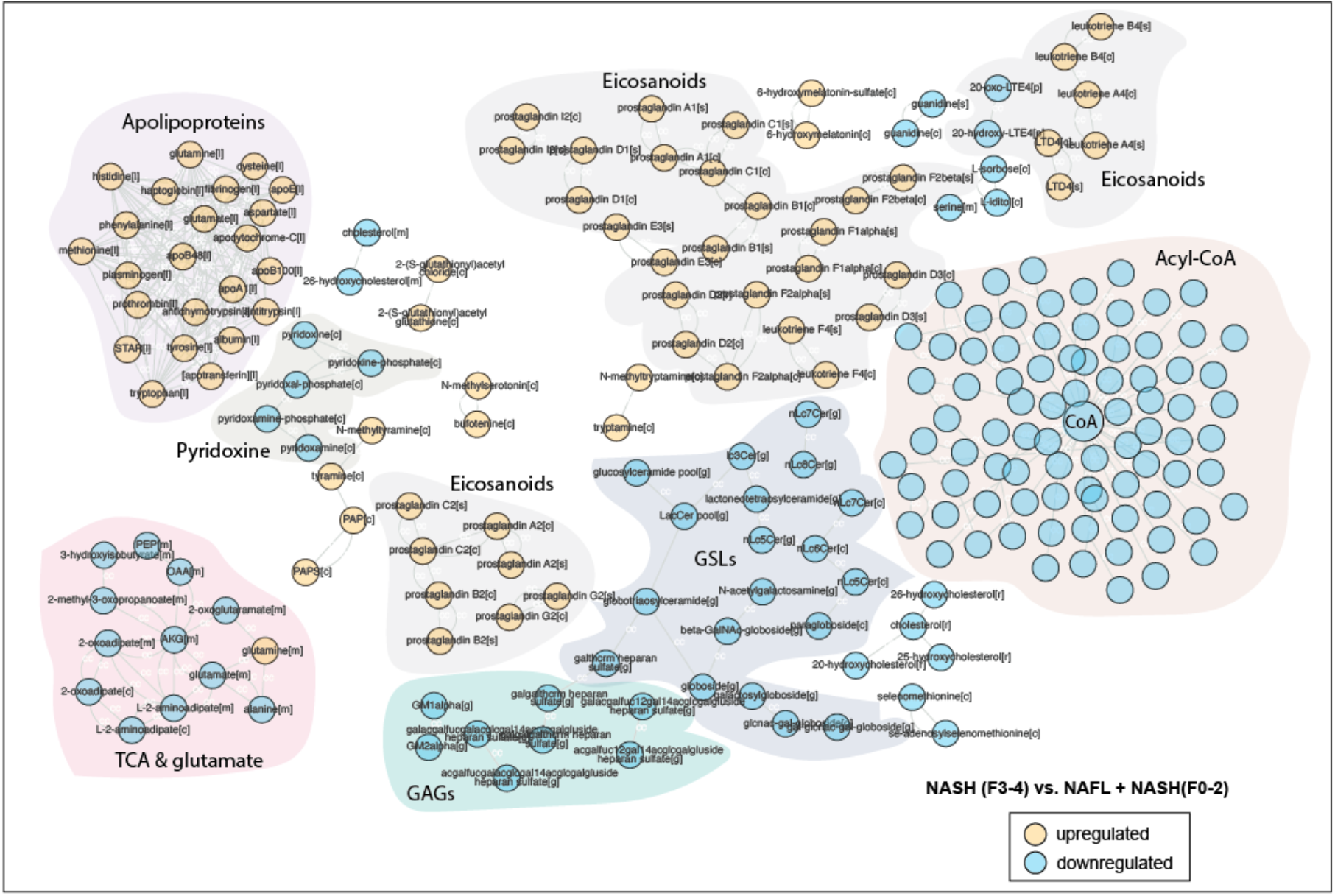
A metabolic-centric view of reporter metabolite (RM) modules that were significantly altered between ‘advanced’ fibrosis *vs*. ‘mild disease. Orange and blue color denotes up and downregulated (p<0.05 corrected for FDR) respectively. Each node represents a ‘RM’ and single or double lines represent reversible or irreversible metabolic reactions respectively. RMs that belong to a particular chemical class are color coded.

**Figure S8.**
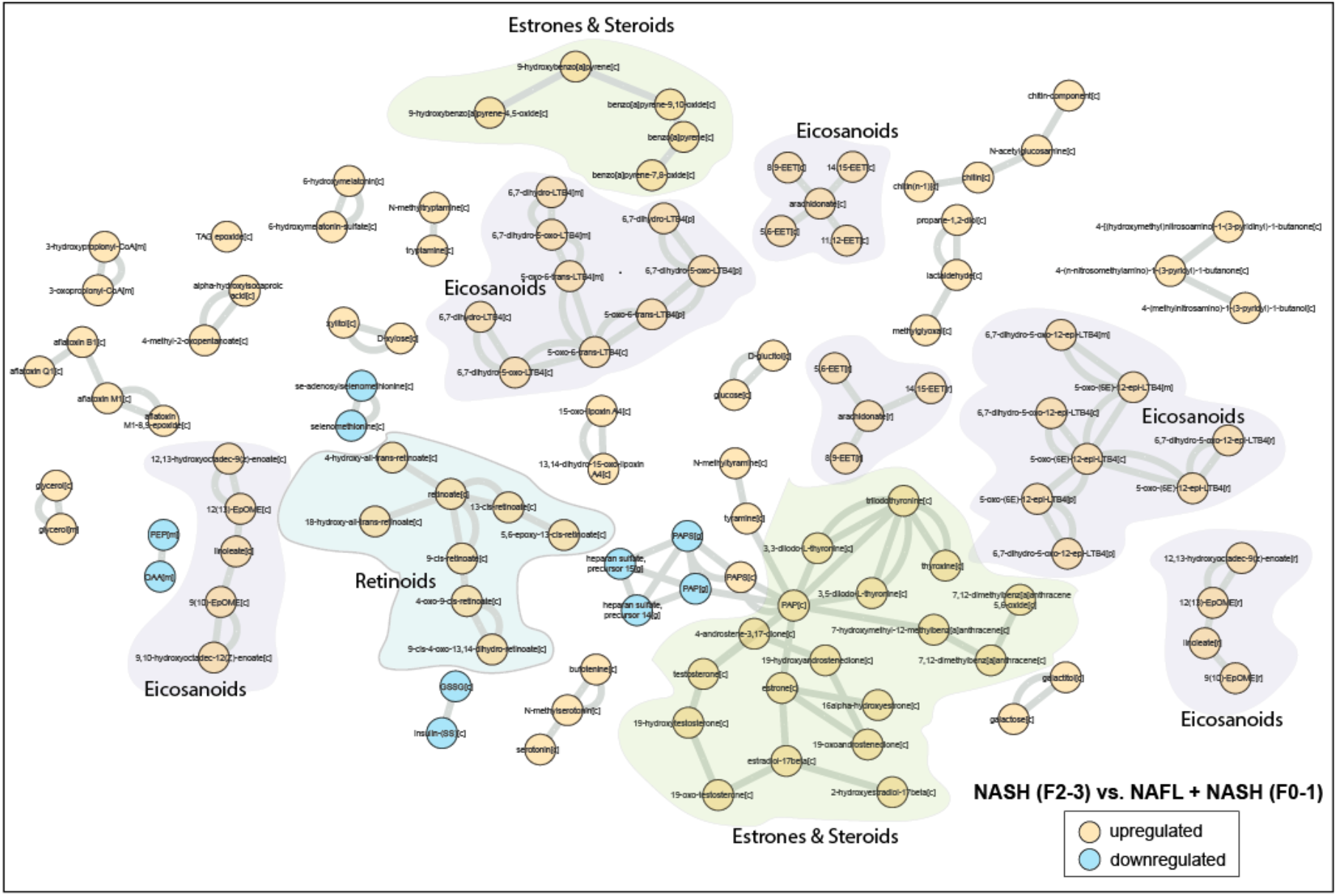
A metabolic-centric view of reporter metabolite (RM) modules that were significantly altered between ‘clinically significant’ non-cirrhotic fibrosis *vs*. ‘minimum’ disease. Orange and blue color denotes up and downregulated (p<0.05 corrected for FDR) respectively. Each node represents a ‘RM’ and single or double lines represent reversible or irreversible metabolic reactions respectively. RMs that belong to a particular chemical class are color coded.

**Figure S9.**
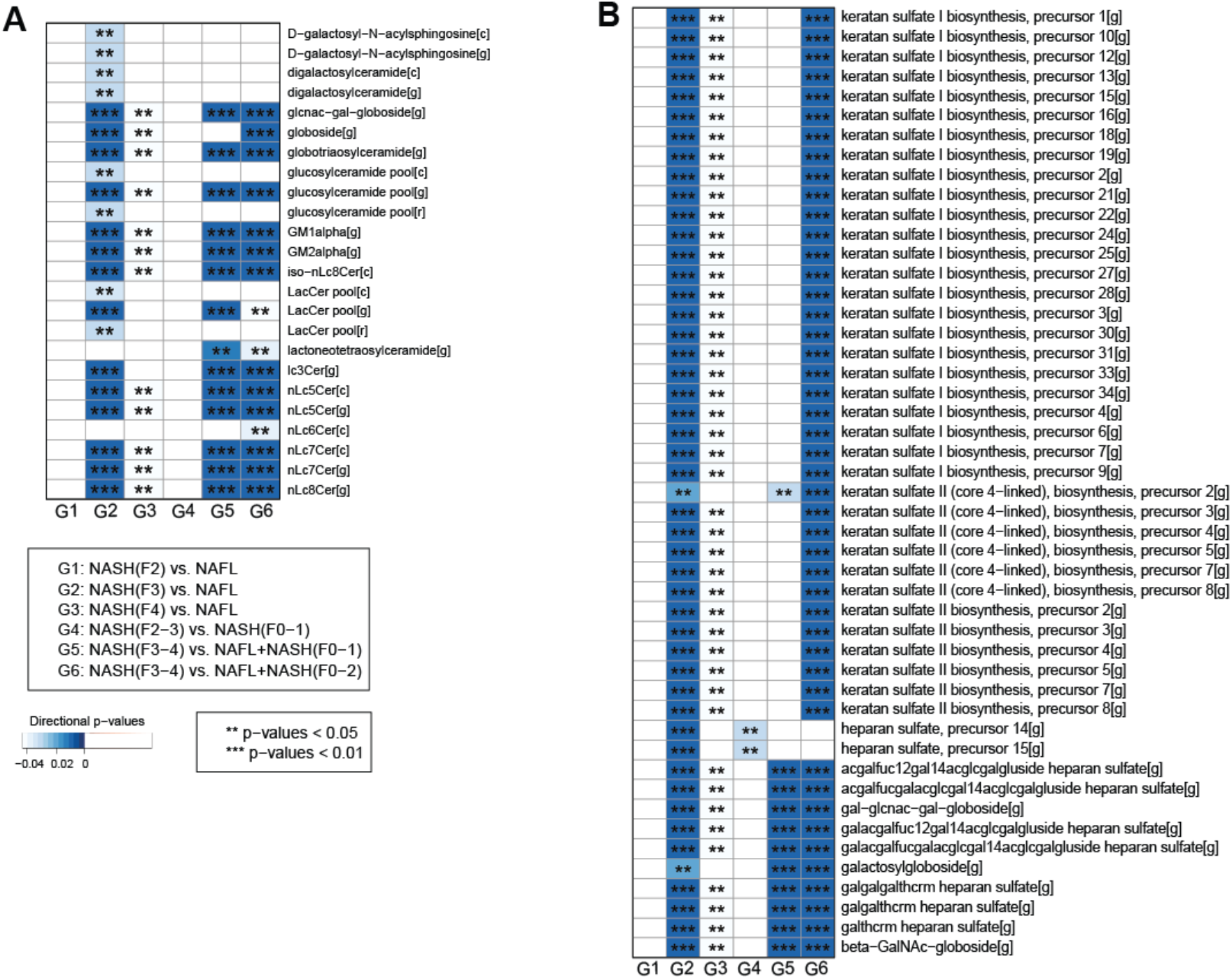
Heatmap showing RM clusters of (**A**) glycosphingolipids (GSLs) and (**B**) glycosaminoglycans (GAGs) that are significantly (***p<0.01, **p<0.05, corrected for FDR) downregulated (blue color) along the different stages NAFLD.

**Figure S10.**
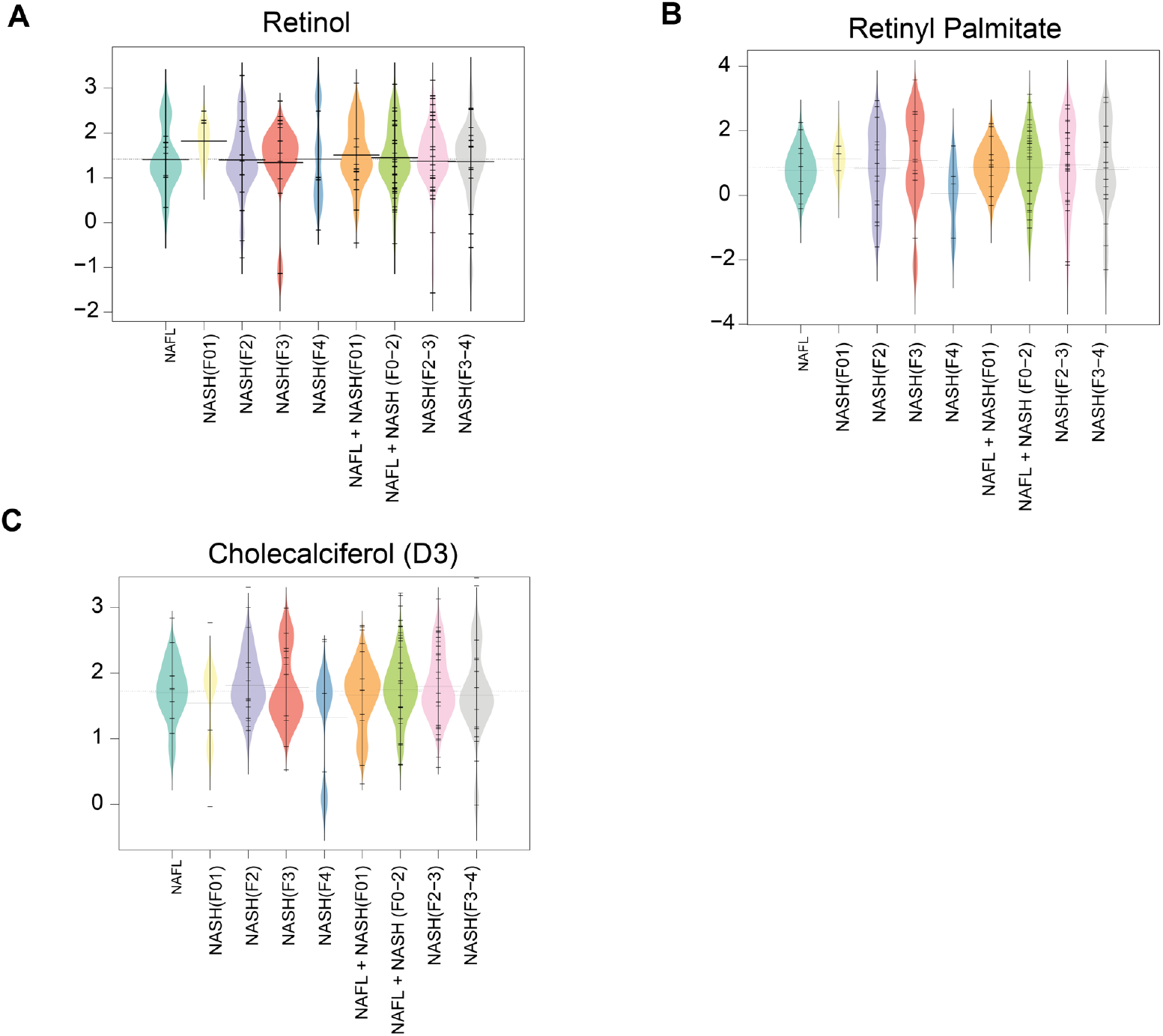
Serum metabolic levels of ceramides, glycosphingolipids and retinoids in the patients at different stages of NAFLD. (**A-C**) Beanplots showing the log_2_ intensities of the metabolites across different stages of NAFLD. The dotted line denotes the mean of the population, and the black dashed lines in the bean plots represent the group mean.

**Figure S11.**
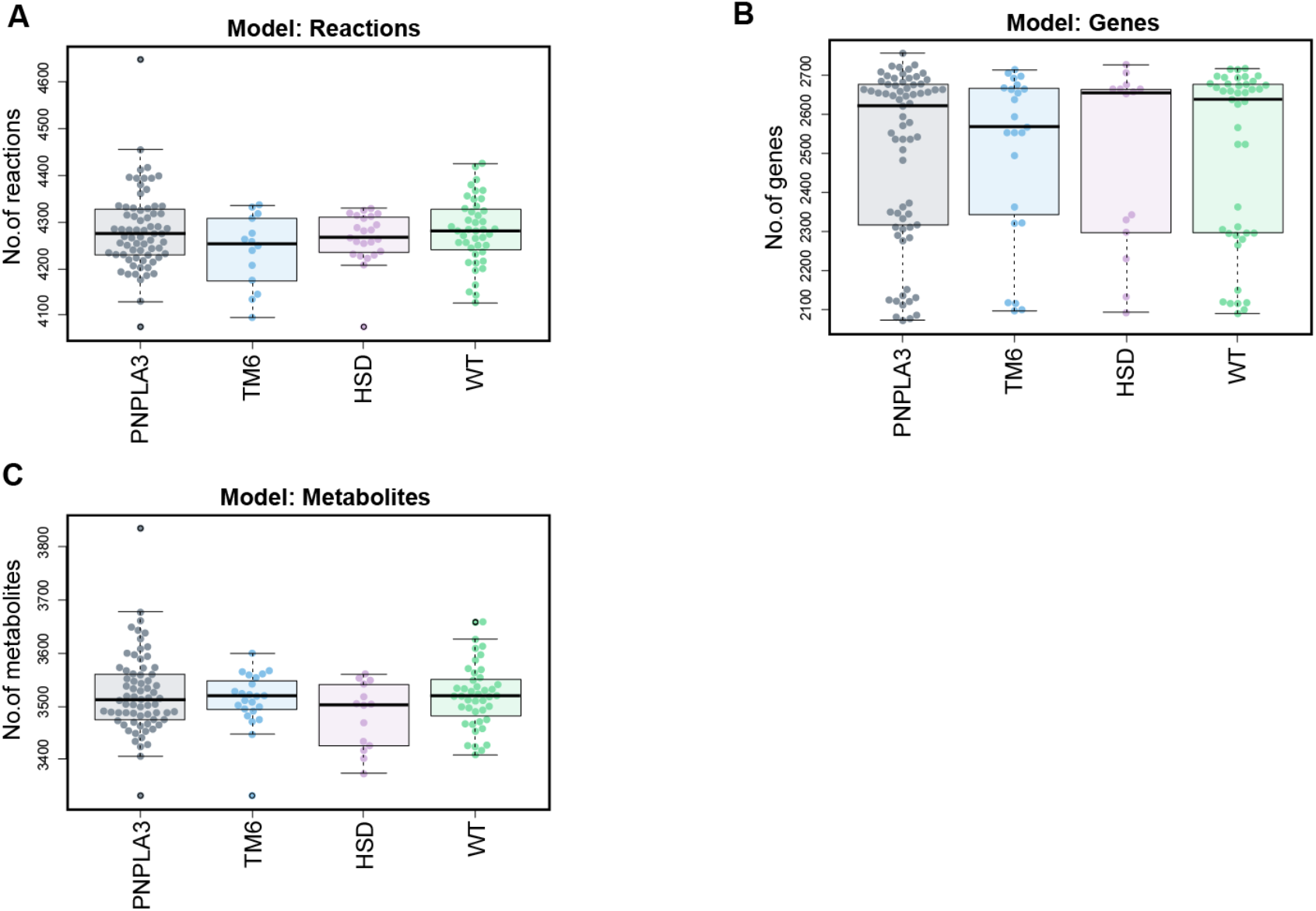
Personalized genome-scale metabolic models of human hepatocytes contextualized for gene variants and WT. Boxplots showing distribution of number of reactions (**A**), genes (**B**) and metabolites (**C**) (marked by dots) contained in the genome-scale metabolic models (GEMs) of human hepatocytes developed for (n=139) subjects, and contextualized for three major genetic variants (*PNPLA3, TM6SF2* and *HSD17B13*) and WT; associated with risk and severity of NAFLD. ‘WT’ is a wild-type model.

**Figure S12.**
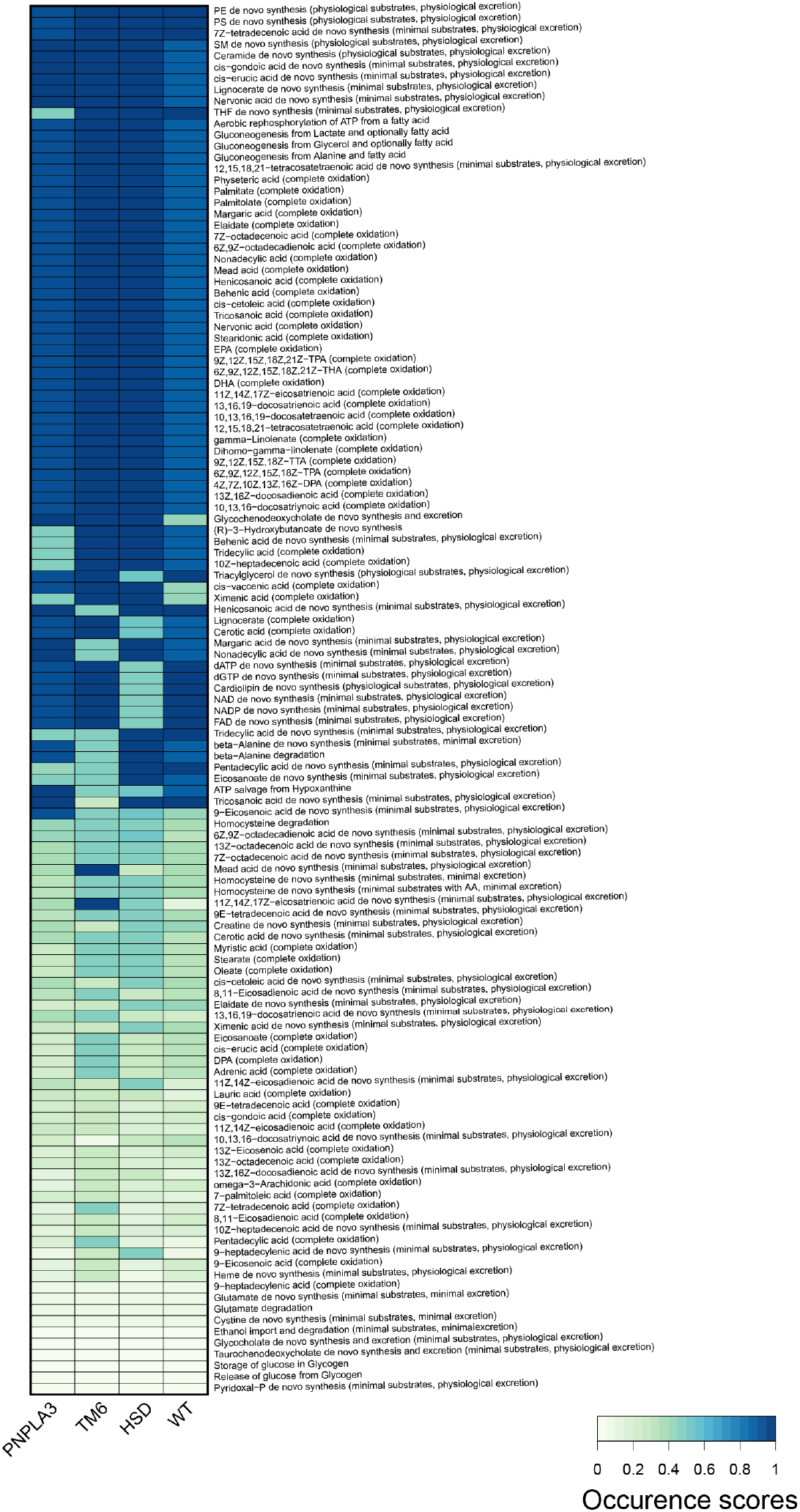
Metabolic tasks carried out by the personalized GEMs contextualized for the gene variants and wild type (WT). [0-1] denotes low to high occurrence scores (OS). OS acquired by a GEM is based on its capability (‘Yes’ or ‘No’) to perform the assigned metabolic task.

**Figure S13.**
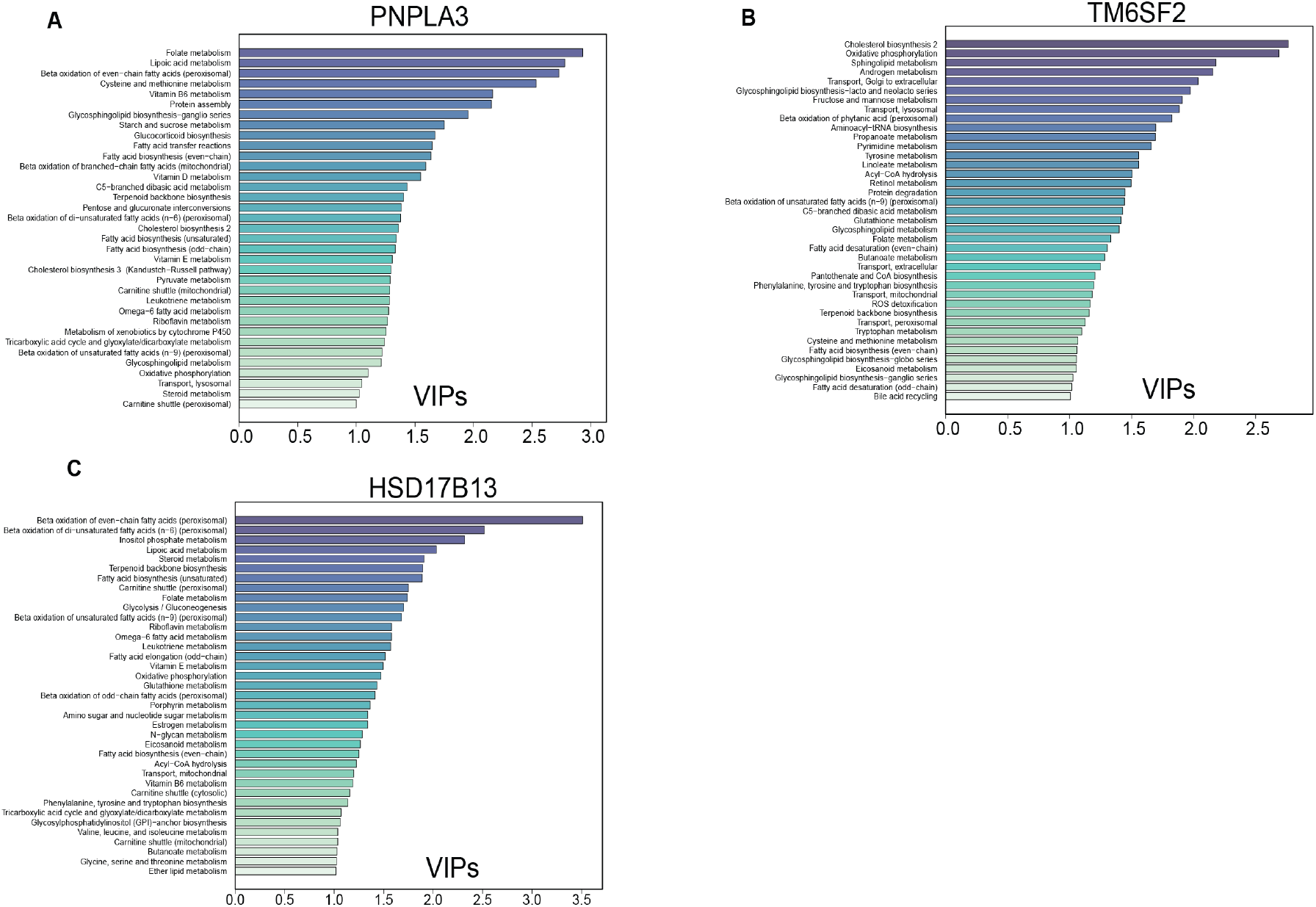
(**A-C**) Metabolic subsystems sorted by their (VIP scores >1) retrieved by fitting pairwise PLS-DA models for (**A**) *PNPLA3* (**B**) *TM6SF2* and (**C**) *HSD17B13* gene variants vs. WT.

**Table S1.**
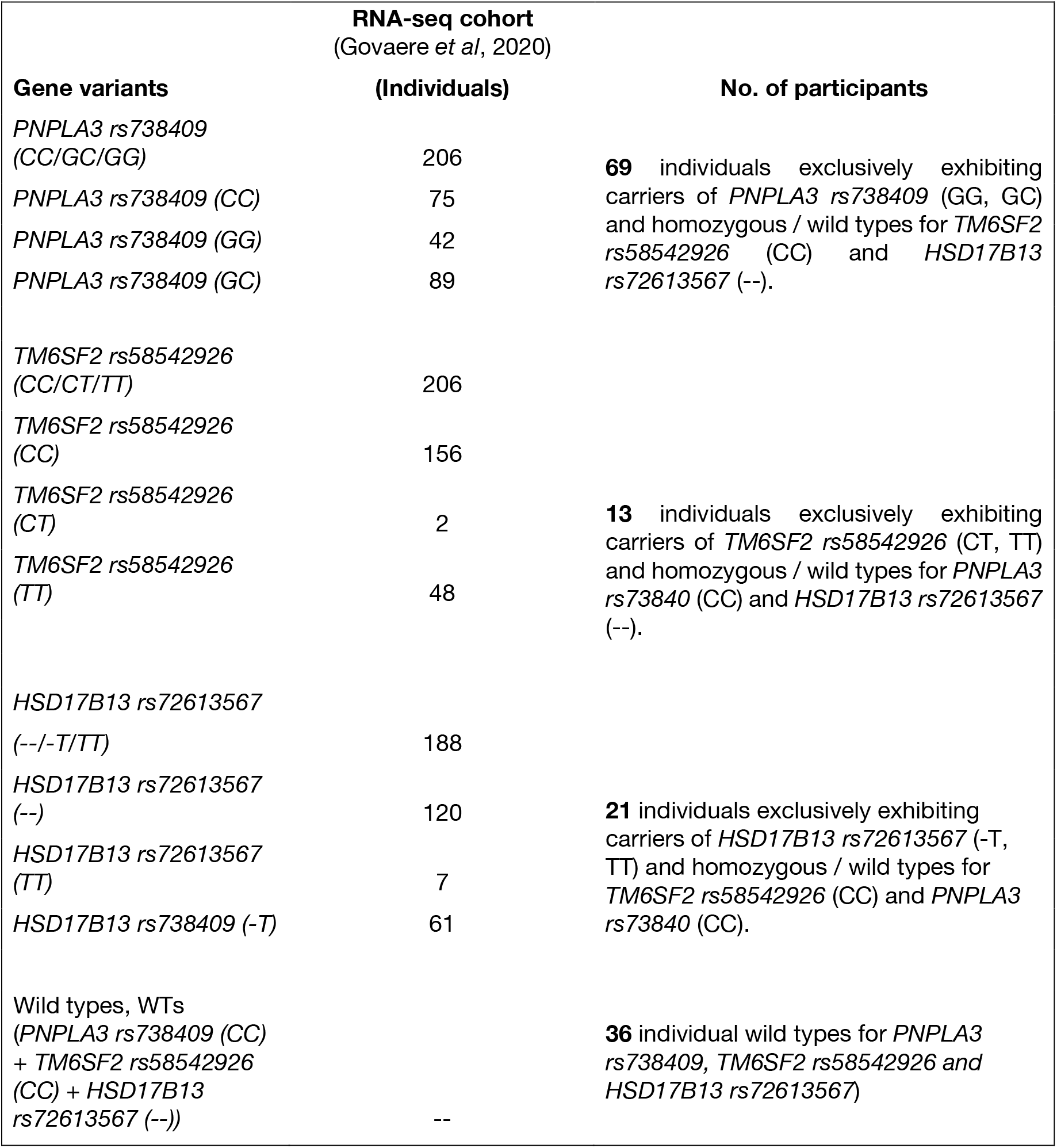
Selection of individuals carrying variants for *PNPLA3, TM6SF2* and *HSD17B13* genes associated with risk and severity of NAFLD.

## References

Abul-Husn, N.S., Cheng, X., Li, A.H., Xin, Y., Schurmann, C., Stevis, P., Liu, Y., Kozlitina, J., Stender, S., Wood, G.C., et al. (2018). A Protein-Truncating HSD17B13 Variant and Protection from Chronic Liver Disease. N Engl J Med 378, 1096–1106.

Agren, R., Bordel, S., Mardinoglu, A., Pornputtapong, N., Nookaew, I., and Nielsen, J. (2012). Reconstruction of genome-scale active metabolic networks for 69 human cell types and 16 cancer types using INIT. PLoS Comput Biol 8, e1002518.

Anstee, Q.M., Darlay, R., Cockell, S., Meroni, M., Govaere, O., Tiniakos, D., Burt, A.D., Bedossa, P., Palmer, J., Liu, Y.L., et al. (2020). Genome-wide association study of non-alcoholic fatty liver and steatohepatitis in a histologically characterised cohort(). J Hepatol 73, 505–515.

Anstee, Q.M., Reeves, H.L., Kotsiliti, E., Govaere, O., and Heikenwalder, M. (2019). From NASH to HCC: current concepts and future challenges. Nat Rev Gastroenterol Hepatol 16, 411–428.

Anstee, Q.M., Targher, G., and Day, C.P. (2013). Progression of NAFLD to diabetes mellitus, cardiovascular disease or cirrhosis. Nat Rev Gastroenterol Hepatol 10, 330–344.

Apostolidis, S.A., Rodríguez-Rodríguez, N., Suárez-Fueyo, A., Dioufa, N., Ozcan, E., Crispín, J.C., Tsokos, M.G., and Tsokos, G.C. (2016). Phosphatase PP2A is requisite for the function of regulatory T cells. Nature immunology 17, 556.

Arendt, B.M., Comelli, E.M., Ma, D.W., Lou, W., Teterina, A., Kim, T., Fung, S.K., Wong, D.K., McGilvray, I., Fischer, S.E., et al. (2015). Altered hepatic gene expression in nonalcoholic fatty liver disease is associated with lower hepatic n-3 and n-6 polyunsaturated fatty acids. Hepatology 61, 1565–1578.

Bordbar, A., Mo, M.L., Nakayasu, E.S., Schrimpe-Rutledge, A.C., Kim, Y.M., Metz, T.O., Jones, M.B., Frank, B.C., Smith, R.D., Peterson, S.N., et al. (2012). Model-driven multi-omic data analysis elucidates metabolic immunomodulators of macrophage activation. Mol Syst Biol 8, 558.

Bordel, S., Agren, R., and Nielsen, J. (2010). Sampling the solution space in genome-scale metabolic networks reveals transcriptional regulation in key enzymes. PLoS Comput Biol 6, e1000859.

Cakir, T., Patil, K.R., Onsan, Z., Ulgen, K.O., Kirdar, B., and Nielsen, J. (2006). Integration of metabolome data with metabolic networks reveals reporter reactions. Mol Syst Biol 2, 50.

Carey, R.N., Wold, S., and Westgard, J.O. (1975). Principal component analysis: an alternative to “referee” methods in method comparison studies. Anal Chem 47, 1824–1829.

Castillo, S., Mattila, I., Miettinen, J., Oresic, M., and Hyotylainen, T. (2011). Data Analysis Tool for Comprehensive Two-Dimensional Gas Chromatography/Time-of-Flight Mass Spectrometry. Analytical Chemistry 83, 3058–3067.

Colijn, C., Brandes, A., Zucker, J., Lun, D.S., Weiner, B., Farhat, M.R., Cheng, T.-Y., Moody, D.B., Murray, M., and Galagan, J.E. (2009). Interpreting expression data with metabolic flux models: predicting Mycobacterium tuberculosis mycolic acid production. PLoS computational biology 5, e1000489.

Estes, C., Anstee, Q.M., Arias-Loste, M.T., Bantel, H., Bellentani, S., Caballeria, J., Colombo, M., Craxi, A., Crespo, J., Day, C.P., et al. (2018). Modeling NAFLD disease burden in China, France, Germany, Italy, Japan, Spain, United Kingdom, and United States for the period 2016-2030. J Hepatol 69, 896–904.

Farrés, M., Platikanov, S., Tsakovski, S., and Tauler, R. (2015). Comparison of the variable importance in projection (VIP) and of the selectivity ratio (SR) methods for variable selection and interpretation. J Chemom 29, 528–536.

Gault, C.R., Obeid, L.M., and Hannun, Y.A. (2010). An overview of sphingolipid metabolism: from synthesis to breakdown. Adv Exp Med Biol 688, 1–23.

Govaere, O., Cockell, S., Tiniakos, D., Queen, R., Younes, R., Vacca, M., Alexander, L., Ravaioli, F., Palmer, J., Petta, S., et al. (2020). Transcriptomic profiling across the nonalcoholic fatty liver disease spectrum reveals gene signatures for steatohepatitis and fibrosis. Sci Transl Med 12, eaba4448.

Haas, J.T., Vonghia, L., Mogilenko, D.A., Verrijken, A., Molendi-Coste, O., Fleury, S., Deprince, A., Nikitin, A., Woitrain, E., Ducrocq-Geoffroy, L., et al. (2019). Transcriptional Network Analysis Implicates Altered Hepatic Immune Function in NASH development and resolution. Nat Metab 1, 604–614.

Hardy, T., Wonders, K., Younes, R., Aithal, G.P., Aller, R., Allison, M., Bedossa, P., Betsou, F., Boursier, J., Brosnan, M.J., et al. (2020). The European NAFLD Registry: A real-world longitudinal cohort study of nonalcoholic fatty liver disease. Contemp Clin Trials 98, 106175.

He, S., McPhaul, C., Li, J.Z., Garuti, R., Kinch, L., Grishin, N.V., Cohen, J.C., and Hobbs, H.H. (2010). A sequence variation (I148M) in PNPLA3 associated with nonalcoholic fatty liver disease disrupts triglyceride hydrolysis. J Biol Chem 285, 6706–6715.

Heirendt, L., Arreckx, S., Pfau, T., Mendoza, S.N., Richelle, A., Heinken, A., Haraldsdottir, H.S., Keating, S.M., Vlasov, V., and Wachowiak, J. (2017). Creation and analysis of biochemical constraint-based models: the COBRA Toolbox v3. 0. arXiv preprint 171004038.

Herrmann, H.A., Dyson, B.C., Vass, L., Johnson, G.N., and Schwartz, J.M. (2019). Flux sampling is a powerful tool to study metabolism under changing environmental conditions. NPJ Syst Biol Appl 5, 32.

Hyotylainen, T., Jerby, L., Petaja, E.M., Mattila, I., Jantti, S., Auvinen, P., Gastaldelli, A., Yki-Jarvinen, H., Ruppin, E., and Oresic, M. (2016). Genome-scale study reveals reduced metabolic adaptability in patients with non-alcoholic fatty liver disease. Nat Commun 7, 8994.

Ishay, Y., Nachman, D., Khoury, T., and Ilan, Y. (2020). The role of the sphingolipid pathway in liver fibrosis: an emerging new potential target for novel therapies. Am J Physiol Cell Physiol 318, C1055–C1064.

Jerby, L., Shlomi, T., and Ruppin, E. (2010). Computational reconstruction of tissue-specific metabolic models: application to human liver metabolism. Mol Syst Biol 6, 401.

Kleiner, D.E., Brunt, E.M., Van Natta, M., Behling, C., Contos, M.J., Cummings, O.W., Ferrell, L.D., Liu, Y.C., Torbenson, M.S., Unalp-Arida, A., et al. (2005). Design and validation of a histological scoring system for nonalcoholic fatty liver disease. Hepatology 41, 1313–1321.

Kotronen, A., Seppanen-Laakso, T., Westerbacka, J., Kiviluoto, T., Arola, J., Ruskeepaa, A.-L., Oresic, M., and Yki-Jarvinen, H. (2009). Hepatic Stearoyl-CoA Desaturase (SCD)-1 Activity and Diacylglycerol but Not Ceramide Concentrations Are Increased in the Nonalcoholic Human Fatty Liver. Diabetes 58, 203–208.

Kovarova, M., Konigsrainer, I., Konigsrainer, A., Machicao, F., Haring, H.U., Schleicher, E., and Peter, A. (2015). The Genetic Variant I148M in PNPLA3 Is Associated With Increased Hepatic Retinyl-Palmitate Storage in Humans. J Clin Endocrinol Metab 100, E1568–1574.

Labenz, C., Huber, Y., Kalliga, E., Nagel, M., Ruckes, C., Straub, B.K., Galle, P.R., Worns, M.A., Anstee, Q.M., Schuppan, D., et al. (2018). Predictors of advanced fibrosis in non-cirrhotic non-alcoholic fatty liver disease in Germany. Aliment Pharmacol Ther 48, 1109–1116.

Le Cao, K.A., Boitard, S., and Besse, P. (2011). Sparse PLS discriminant analysis: biologically relevant feature selection and graphical displays for multiclass problems. BMC Bioinformatics 12, 253.

Lefebvre, P., Lalloyer, F., Bauge, E., Pawlak, M., Gheeraert, C., Dehondt, H., Vanhoutte, J., Woitrain, E., Hennuyer, N., Mazuy, C., et al. (2017). Interspecies NASH disease activity whole-genome profiling identifies a fibrogenic role of PPARalpha-regulated dermatopontin. JCI Insight 2, e92264.

Liu, Y.L., Reeves, H.L., Burt, A.D., Tiniakos, D., McPherson, S., Leathart, J.B., Allison, M.E., Alexander, G.J., Piguet, A.C., Anty, R., et al. (2014). TM6SF2 rs58542926 influences hepatic fibrosis progression in patients with non-alcoholic fatty liver disease. Nat Commun 5, 4309.

Luukkonen, P.K., Zhou, Y., Nidhina Haridas, P.A., Dwivedi, O.P., Hyotylainen, T., Ali, A., Juuti, A., Leivonen, M., Tukiainen, T., Ahonen, L., et al. (2017). Impaired hepatic lipid synthesis from polyunsaturated fatty acids in TM6SF2 E167K variant carriers with NAFLD. J Hepatol 67, 128–136.

Ma, Y., Belyaeva, O.V., Brown, P.M., Fujita, K., Valles, K., Karki, S., de Boer, Y.S., Koh, C., Chen, Y., Du, X., et al. (2019). 17-Beta Hydroxysteroid Dehydrogenase 13 Is a Hepatic Retinol Dehydrogenase Associated With Histological Features of Nonalcoholic Fatty Liver Disease. Hepatology 69, 1504–1519.

Mardinoglu, A., Agren, R., Kampf, C., Asplund, A., Nookaew, I., Jacobson, P., Walley, A.J., Froguel, P., Carlsson, L.M., Uhlen, M., et al. (2013). Integration of clinical data with a genome-scale metabolic model of the human adipocyte. Mol Syst Biol 9, 649.

Mardinoglu, A., Agren, R., Kampf, C., Asplund, A., Uhlen, M., and Nielsen, J. (2014). Genome-scale metabolic modelling of hepatocytes reveals serine deficiency in patients with non-alcoholic fatty liver disease. Nat Commun 5, 3083.

Masoodi, M., Gastaldelli, A., Hyötyläinen, T., Arretxe, E., Alonso, C., Gaggini, M., Brosnan, J., Anstee, Q.M., Millet, O., Ortiz, P., et al. (2021, in press). Metabolomics and lipidomics in NAFLD: biomarkers and non-invasive diagnostic tests. Nat Rev Gastroenterol Hepatol, 10.1038/s41575-41021-00502-41579.

McPherson, S., Hardy, T., Henderson, E., Burt, A.D., Day, C.P., and Anstee, Q.M. (2015). Evidence of NAFLD progression from steatosis to fibrosing-steatohepatitis using paired biopsies: Implications for prognosis and clinical management. J Hepatol 62, 1148–1155.

Mondul, A., Mancina, R.M., Merlo, A., Dongiovanni, P., Rametta, R., Montalcini, T., Valenti, L., Albanes, D., and Romeo, S. (2015). PNPLA3 I148M Variant Influences Circulating Retinol in Adults with Nonalcoholic Fatty Liver Disease or Obesity. J Nutr 145, 1687–1691.

Moylan, C.A., Pang, H., Dellinger, A., Suzuki, A., Garrett, M.E., Guy, C.D., Murphy, S.K., Ashley-Koch, A.E., Choi, S.S., Michelotti, G.A., et al. (2014). Hepatic gene expression profiles differentiate presymptomatic patients with mild versus severe nonalcoholic fatty liver disease. Hepatology 59, 471–482.

Nygren, H., Seppanen-Laakso, T., Castillo, S., Hyotylainen, T., and Oresic, M. (2011). Liquid chromatography-mass spectrometry (LC-MS)-based lipidomics for studies of body fluids and tissues. Methods Mol Biol 708, 247–257.

Opdam, S., Richelle, A., Kellman, B., Li, S., Zielinski, D.C., and Lewis, N.E. (2017). A Systematic Evaluation of Methods for Tailoring Genome-Scale Metabolic Models. Cell Syst 4, 318–329 e316.

Oresic, M., Hyotylainen, T., Kotronen, A., Gopalacharyulu, P., Nygren, H., Arola, J., Castillo, S., Mattila, I., Hakkarainen, A., Borra, R.J.H., et al. (2013). Prediction of non-alcoholic fatty-liver disease and liver fat content by serum molecular lipids. Diabetologia 56, 2266–2274.

Osterlund, T., Nookaew, I., Bordel, S., and Nielsen, J. (2013). Mapping condition-dependent regulation of metabolism in yeast through genome-scale modeling. BMC Syst Biol 7, 36.

Pagadala, M., Kasumov, T., McCullough, A.J., Zein, N.N., and Kirwan, J.P. (2012). Role of ceramides in nonalcoholic fatty liver disease. Trends Endocrinol Metab 23, 365–371.

Patil, K.R., and Nielsen, J. (2005). Uncovering transcriptional regulation of metabolism by using metabolic network topology. Proceedings of the National Academy of Sciences of the United States of America 102, 2685–2689.

Pettinelli, P., Arendt, B.M., Teterina, A., McGilvray, I., Comelli, E.M., Fung, S.K., Fischer, S.E., and Allard, J.P. (2018). Altered hepatic genes related to retinol metabolism and plasma retinol in patients with non-alcoholic fatty liver disease. PLoS One 13, e0205747.

Pluskal, T., Castillo, S., Villar-Briones, A., and Oresic, M. (2010). MZmine 2: modular framework for processing, visualizing, and analyzing mass spectrometry-based molecular profile data. BMC Bioinformatics 11, 395.

R Development Core Team (2018). R: A language and environment for statistical computing (Vienna: R Foundation for Statistical Computing).

Romeo, S., Kozlitina, J., Xing, C., Pertsemlidis, A., Cox, D., Pennacchio, L.A., Boerwinkle, E., Cohen, J.C., and Hobbs, H.H. (2008). Genetic variation in PNPLA3 confers susceptibility to nonalcoholic fatty liver disease. Nat Genet 40, 1461–1465.

Saeed, A., Dullaart, R.P.F., Schreuder, T., Blokzijl, H., and Faber, K.N. (2017). Disturbed Vitamin A Metabolism in Non-Alcoholic Fatty Liver Disease (NAFLD). Nutrients 10, 29.

Sanyal, A.J., Chalasani, N., Kowdley, K.V., McCullough, A., Diehl, A.M., Bass, N.M., Neuschwander-Tetri, B.A., Lavine, J.E., Tonascia, J., Unalp, A., et al. (2010). Pioglitazone, vitamin E, or placebo for nonalcoholic steatohepatitis. N Engl J Med 362, 1675–1685.

Sen, P., Dickens, A.M., Lopez-Bascon, M.A., Lindeman, T., Kemppainen, E., Lamichhane, S., Ronkko, T., Ilonen, J., Toppari, J., Veijola, R., et al. (2020). Metabolic alterations in immune cells associate with progression to type 1 diabetes. Diabetologia 63, 1017–1031.

Sen, P., Hyotylainen, T., and Oresic, M. (2021). 1-Deoxyceramides - Key players in lipotoxicity and progression to type 2 diabetes? Acta Physiol (Oxf) 232, e13635.

Sen, P., Kemppainen, E., and Orešič, M. (2017). Perspectives on Systems Modelling of Human Peripheral Blood Mononuclear Cells. Frontiers in molecular biosciences 4, 96.

Starmann, J., Falth, M., Spindelbock, W., Lanz, K.L., Lackner, C., Zatloukal, K., Trauner, M., and Sultmann, H. (2012). Gene expression profiling unravels cancer-related hepatic molecular signatures in steatohepatitis but not in steatosis. PLoS One 7, e46584.

Taylor, R.S., Taylor, R.J., Bayliss, S., Hagstrom, H., Nasr, P., Schattenberg, J.M., Ishigami, M., Toyoda, H., Wai-Sun Wong, V., Peleg, N., et al. (2020). Association Between Fibrosis Stage and Outcomes of Patients With Nonalcoholic Fatty Liver Disease: A Systematic Review and Meta-Analysis. Gastroenterology 158, 1611–1625 e1612.

Thiele, I., and Palsson, B.O. (2010). A protocol for generating a high-quality genome-scale metabolic reconstruction. Nat Protoc 5, 93–121.

Trepo, E., and Valenti, L. (2020). Update on NAFLD genetics: From new variants to the clinic. J Hepatol 72, 1196–1209.

Turpin-Nolan, S.M., and Bruning, J.C. (2020). The role of ceramides in metabolic disorders: when size and localization matters. Nat Rev Endocrinol 16, 224–233.

Wang, H., Marcisauskas, S., Sanchez, B.J., Domenzain, I., Hermansson, D., Agren, R., Nielsen, J., and Kerkhoven, E.J. (2018). RAVEN 2.0: A versatile toolbox for metabolic network reconstruction and a case study on Streptomyces coelicolor. PLoS Comput Biol 14, e1006541.

Westerbacka, J., Kotronen, A., Fielding, B.A., Wahren, J., Hodson, L., Perttila, J., Seppanen-Laakso, T., Suortti, T., Arola, J., Hultcrantz, R., et al. (2010). Splanchnic Balance of Free Fatty Acids, Endocannabinoids, and Lipids in Subjects With Nonalcoholic Fatty Liver Disease. Gastroenterology 139, 1961–U1232.

Westerhuis, J.A., Hoefsloot, H.C., Smit, S., Vis, D.J., Smilde, A.K., van Velzen, E.J., van Duijnhoven, J.P., and van Dorsten, F.A. (2008). Assessment of PLSDA cross validation. Metabolomics 4, 81–89.

Younossi, Z., Anstee, Q.M., Marietti, M., Hardy, T., Henry, L., Eslam, M., George, J., and Bugianesi, E. (2018). Global burden of NAFLD and NASH: trends, predictions, risk factors and prevention. Nat Rev Gastroenterol Hepatol 15, 11–20.

Zhang, T., de Waard, A.A., Wuhrer, M., and Spaapen, R.M. (2019). The Role of Glycosphingolipids in Immune Cell Functions. Front Immunol 10, 90.

Zhao, H., Przybylska, M., Wu, I.H., Zhang, J., Maniatis, P., Pacheco, J., Piepenhagen, P., Copeland, D., Arbeeny, C., Shayman, J.A., et al. (2009). Inhibiting glycosphingolipid synthesis ameliorates hepatic steatosis in obese mice. Hepatology 50, 85–93.

Zur, H., Ruppin, E., and Shlomi, T. (2010). iMAT: an integrative metabolic analysis tool. Bioinformatics 26, 3140–3142.

## References

Govaere O, Cockell S, Tiniakos D, Queen R, Younes R, Vacca M, Alexander L, Ravaioli F, Palmer J, Petta S et al (2020) Transcriptomic profiling across the nonalcoholic fatty liver disease spectrum reveals gene signatures for steatohepatitis and fibrosis. Sci Transl Med 12: eaba4448

